# Loss of IVNS1ABP, a gigaxonin paralogue, leads to a progeroid neuropathy due to impaired proteostasis

**DOI:** 10.1101/2024.10.23.24315364

**Authors:** Carine Bonnard, Ain Nur Ali, Fang Yuan, Kiat Y. Tan, Yan T. Lim, Puck W. Chan, Nanthini Ramanathan, Nur A.B.M. Kamsani, Vindhya Chaganty, Ding Xiong, Ye S. Tan, Shu-Min Chou, Marwa Chourabi, Gunaseelan Narayanan, Alvin Y.J. Ng, Sumanty Tohari, Meah W. Yang, Birsen Karaman, Gözde T. Turgut, Emmanuelle Szenker-Ravi, Radoslaw M. Sobota, Zhuoyao Chen, Alex N. Bullock, Rajaa Fathallah, Umut Altunoglu, Venkatesh Byrappa, Ajay S. Mathuru, Mohammad Shboul, Su-Chun Zhang, Bruno Reversade

## Abstract

Impaired proteostasis can induce protein aggregation which is toxic to neuronal cells, contributing to neurodegeneration and other signs of aging. In this study, we delineate an early-onset progressive neuropathy evoking Giant Axonal Neuropathy 1. The causative gene *IVNS1ABP* encodes a E3-ubiquitin ligase adaptor which is a close gigaxonin paralogue. Patient-derived fibroblasts, iPSCs, and neural progenitors exhibited hallmarks of protein accumulation and lysosomal dysfunction. Ubiquitome analysis revealed overlapping targets with gigaxonin, including Vimentin and MAP1B. The biallelic correction to the isogenic wildtype state in disease-relevant motor neurons partly rescued cellular vulnerabilities. A newly generated *ivns1abpa/b* knockout zebrafish model partially recapitulated the human peripheral neuropathy, exhibiting aberrant primary motor neuron axon pathfinding, leading to impaired locomotion. Our findings indicate that IVNS1ABP functions in the same pathway as gigaxonin, ensuring appropriate cellular turnover of critical protein substrates, whose accumulation leads to accelerated aging in discrete cellular lineages.

## Introduction

Aging, an irreversible process affecting all humans, is characterized by a gradual decline in homeostasis due to both internal (intrinsic) and external (extrinsic) factors^1,2^. It involves alterations in the capacity to sustain organ function driven by an accumulation of cellular damage^3,4^. As a result, tissue function deteriorates, leading to a substantial increase in aging-related disorders. These disorders encompass cardiovascular diseases, metabolic dysfunctions, autoimmune conditions, musculoskeletal impairments, and neurodegenerative diseases^5,6^.

Aging-related neurological disorders affect the central and peripheral nervous systems, resulting in a range of symptoms including paralysis, muscle weakness, poor coordination, loss of sensation, seizures, confusion, and dementia. The underlying molecular and cellular processes that cause neurodegeneration are classified in a set of hallmarks^7,8^, including 1) genomic instability where DNA repair mechanism loses its efficiency^9^, 2) epigenetic alterations in DNA methylation, chromatin structure and histone modifications, impacting the regulation of gene expression^10,11^, 3) RNA mis-splicing that changes gene expression^12^, 4) mitochondrial malfunction, altering energy homeostasis^13^, 5) cell senescence that leads to inflammation^14,15^, and 6) loss of proteostasis, causing accumulation of protein aggregates that are toxic for the cells^16,17^.

Proteostasis is maintained by cellular processes which control folding, localization, and interactions of proteins from their nascent synthesis through their targeted degradation, the integrity of which is often challenged during aging. Protein accumulation, perturbed protein turnover, ubiquitination modification caused by non-functional ubiquitin proteasome pathway (UPP) are hallmarks of aging and diseases^18–20^. Within the UPP, the chaperone network and autophagy are the primary protein degradation systems that regulate proteostasis. Protein aggregates are removed by a selective autophagy pathway, also termed aggrephagy^21^. The decline or disruption of the aggrephagy pathways results in accumulation of protein aggregates and is associated with a wide range of neurodegenerative disorders^16,22,23^.

Neurodegeneration, as is witnessed in chronological aging, can also be driven by genetic factors^24^. Inherited monogenic variants or risk loci have been identified in patients presenting with premature aging syndromes with clinical features resembling accelerated aging, including early onset neuropathies^25,26^. The investigation of premature aging syndromes has shed light into new pathogenic mechanisms and provides invaluable insights into the broader biology of aging^27,28^. For instance, the identification of deleterious mutations in the PYCR1 and ALDH18 enzymes in patients with progeroid features has highlighted the importance of proline metabolism in aging of connective tissues^29,30^. A large body of work on the rare premature aging disease, Hutchinson-Gilford Progeria Syndrome, demonstrated the role of Lamin A in controlling DNA damage, telomere shortening, genomic instability and senescence, all of which are linked to the hallmarks of aging^31^.

Following this line of investigation, we have sought to investigate patients with severe hereditary neurological diseases that could help us gain inroads into more common neurodegenerative disorders. We recruited three affected patients who display accelerated aging phenotypes, including progressive and degenerative neuropathy accompanied by premature hair whitening. Through homozygosity mapping and exome sequencing, we identified a likely causative biallelic variant in a poorly studied gene *IVNS1ABP*. The encoded IVNS1ABP protein belongs to the BTB-Kelch family of E3 ubiquitin ligase adaptors and shares homology with gigaxonin, suggesting a role in the UPP. Using *in vitro* assays with patients’ cells including primary fibroblasts, iPSCs, NPCs, and *in vivo* disease modeling in zebrafish, our study characterizes the molecular function of IVNS1ABP towards cellular proteostasis. We document that IVNS1ABP is required for motor neuron development and function and possesses overlapping substrates with gigaxonin. We propose that both E3 ligase adaptor proteins work in concert to engage the UPP pathway to clear the accumulation of proteins that would otherwise become toxic and drive neurodegeneration.

## Results

### A homozygous variant in *IVNS1ABP* segregates with a progeroid neuropathy

We investigated a consanguineous family in which three affected patients exhibit an early-onset and progressive neuropathy (Table 1 and Figures 1A and 1B). The proposita II.1, II.4 and II.5 display peripheral neuropathy leading to steppage gait with progressive loss of deambulation (Figure S1A). Patient II.1 walked at 1 year old, but showed signs of regression at 5-10 years old, with difficulty climbing the stairs at 5-10 years old, and lost gait at the age of 10-15. Patients II.4 and II.5 walked at 1.5 years old and showed abnormal gait from 5-10 years old (Movie 1). Skin dyschromatosis with hypo- and hyper-pigmented macules over the face and body were present, the extent of which increased with age (Figure S1B). A notable premature whitening of hairs, eyelashes and eyebrows was seen in all three affected patients (Figure 1B). Consistent with autosomal recessive inheritance, homozygosity mapping followed by exome sequencing in patient II.1 identified a biallelic germline missense variant in the *IVNS1ABP* gene (MIM609209) on chr1:185274675 (GRCh37/hg19) (Table 2 and Figures 1C and S1C). The private mutation NM_006469.4 c.758T>G, p.(Phe253Cys) fully segregated with the disease, the three patients being homozygous, and the unaffected family members either non-carrier (II.2) or heterozygous (II.3, II.6) (Figure 1A). According to publicly available databases and bioinformatics tools, this private allele p.F253C has never been reported in the population and is predicted to be pathogenic (SIFT=0.01, PolyPhen-2=0.993, M-CAP=0.022) (Table 2). According to gnomAD v4.1, no homozygous damaging variants have been reported for *IVNS1ABP*, and the pLoF observed/expected score is 0.32, suggesting that *IVNS1ABP* is intolerant to loss-of-function mutations. A high Combined Annotation Dependent Depletion (CADD) score of 29.9 further demonstrates that this homozygous variant may be deleterious (Figure 1D).

**Figure 1.**
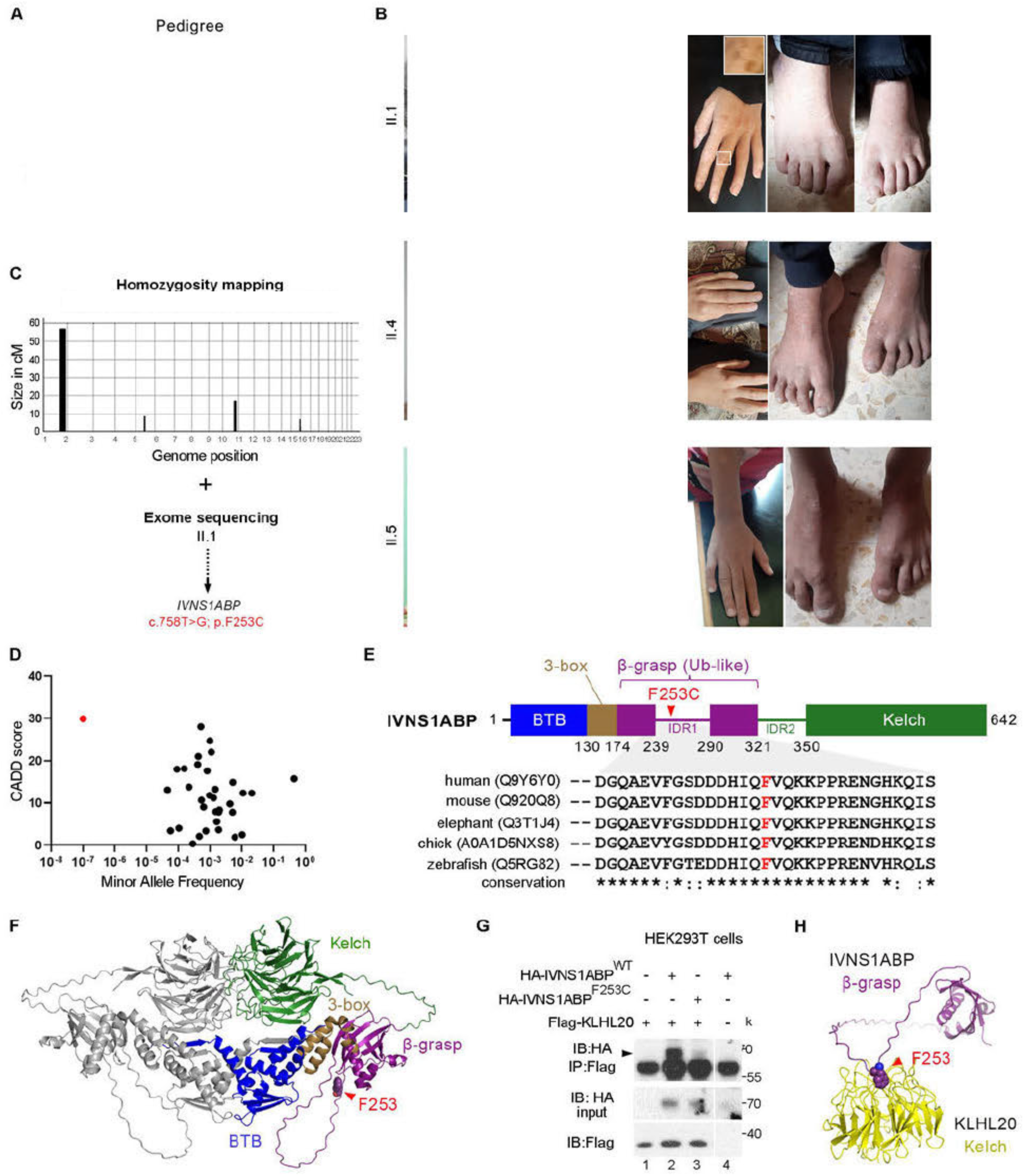
A recessive p.(Phe253Cys) *IVNS1ABP* variant segregates with a segmental syndrome of premature aging **(A)** Pedigree of a consanguineous family with three affected patients carrying a homozygous p.(Phe253Cys) IVNS1ABP variant. Black fill, affected individual; white fill, unaffected individual; green asterisk, skin biopsy; red asterisk, blood drawn. Unaffected family members are either heterozygous (F253C/WT) or non-carrier (WT/WT). Skin biopsies were obtained from unaffected family members (heterozygous I.1, non-carrier II.2) and two affected patients (II.1, II.4). **(B)** Photographs of the three affected patients displaying premature hair whitening and skin dyschromatosis, long fingers, club feet and steppage gait. See also Figures S1A and S1B. **(C)** Homozygosity mapping narrowed down four identical-by-descent (IBD) blocks, and exome sequencing delineated 1 deleterious homozygous (Hmz) variant in IVNS1ABP. See also Figure S1C. **(D)** Minor allele frequency and combined annotation dependent depletion (CADD) score of homozygous *IVNS1ABP* coding variants found in gnomAD v.2.1.1 (black dots) and those found in the affected patients (red dot). **(E)** Domain schematic of the IVNS1ABP protein predicted by Alphafold3, with an N-terminal BTB domain and a C-terminal Kelch domain. In between, a truncated BACK domain with just its 3-box, a ubiquitin-like β-grasp domain, and two intrinsically disordered regions (IDR1 and IDR2), one within the β-grasp and one preceding the Kelch domain. The mutated residue p.F253 (located in IDR1) is highly conserved throughout vertebrate evolution. **(F)** 3D conformation of IVNS1ABP homodimer predicted by Alphafold3. **(G)** Co-immunoprecipitation of transiently overexpressed Ha-tagged wildtype IVNS1ABP^WT^ or mutant IVNS1ABP^F253C^ by Flag-tagged KLHL20, a known IVNS1ABP interacting protein. **(H)** AlphaFold3 prediction of the binding of IVNS1ABP β-grasp fold to KLHL20 Klech domain. See also Figure S1D.

**Table 1.**
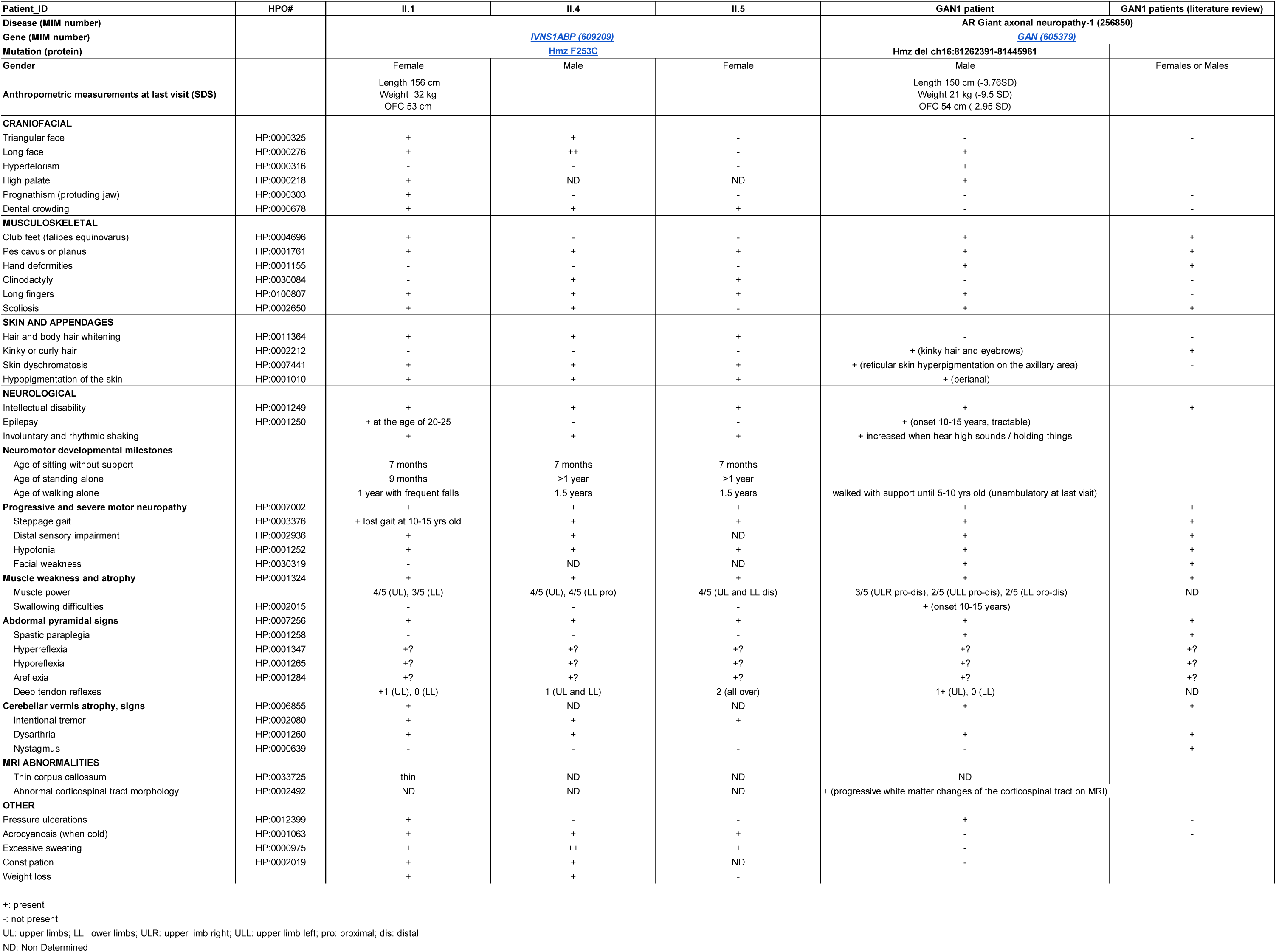
Clinical comparison of patients with biallelic IVNS1ABP variant with recessive GAN1 patients.

**Table 2.**
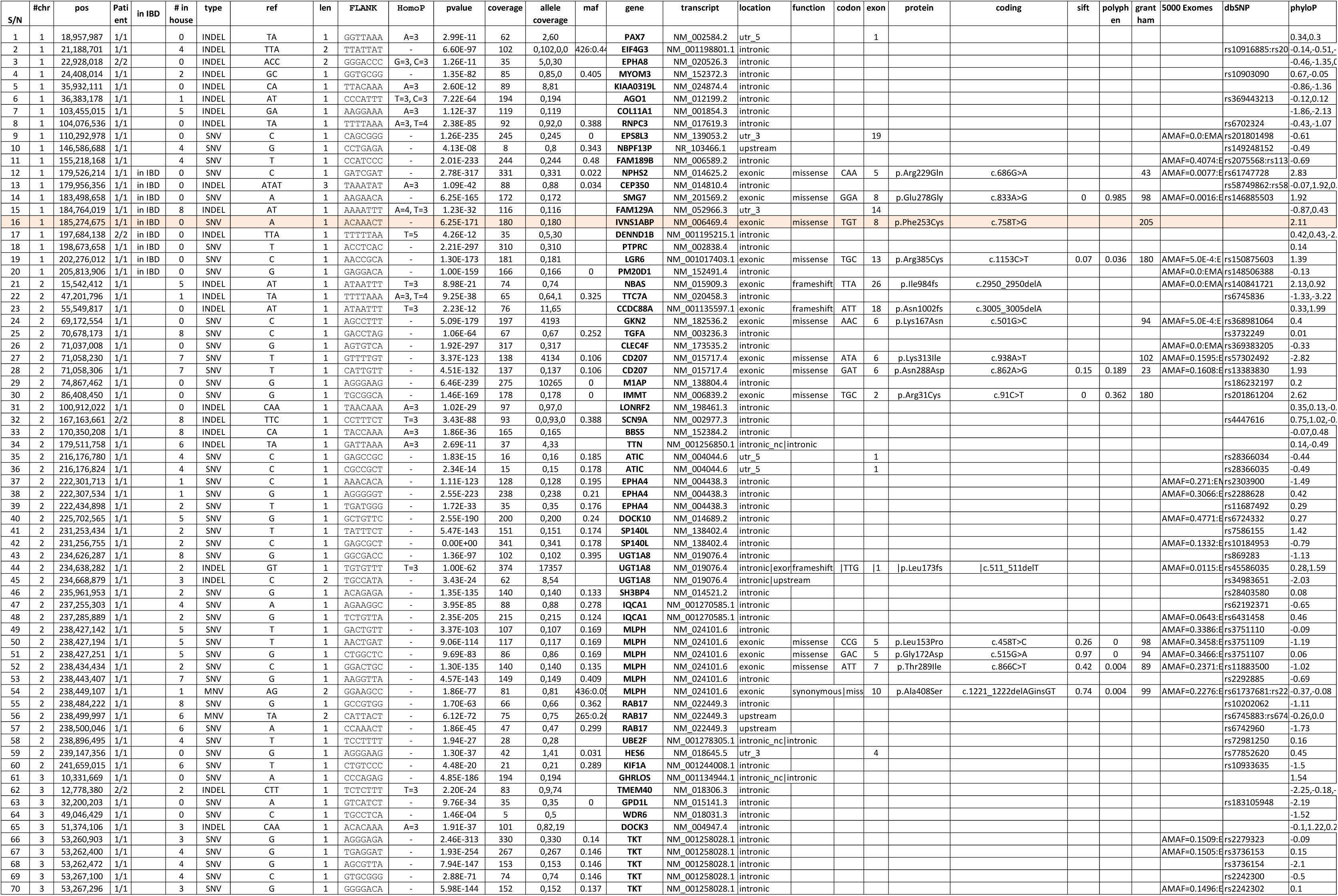
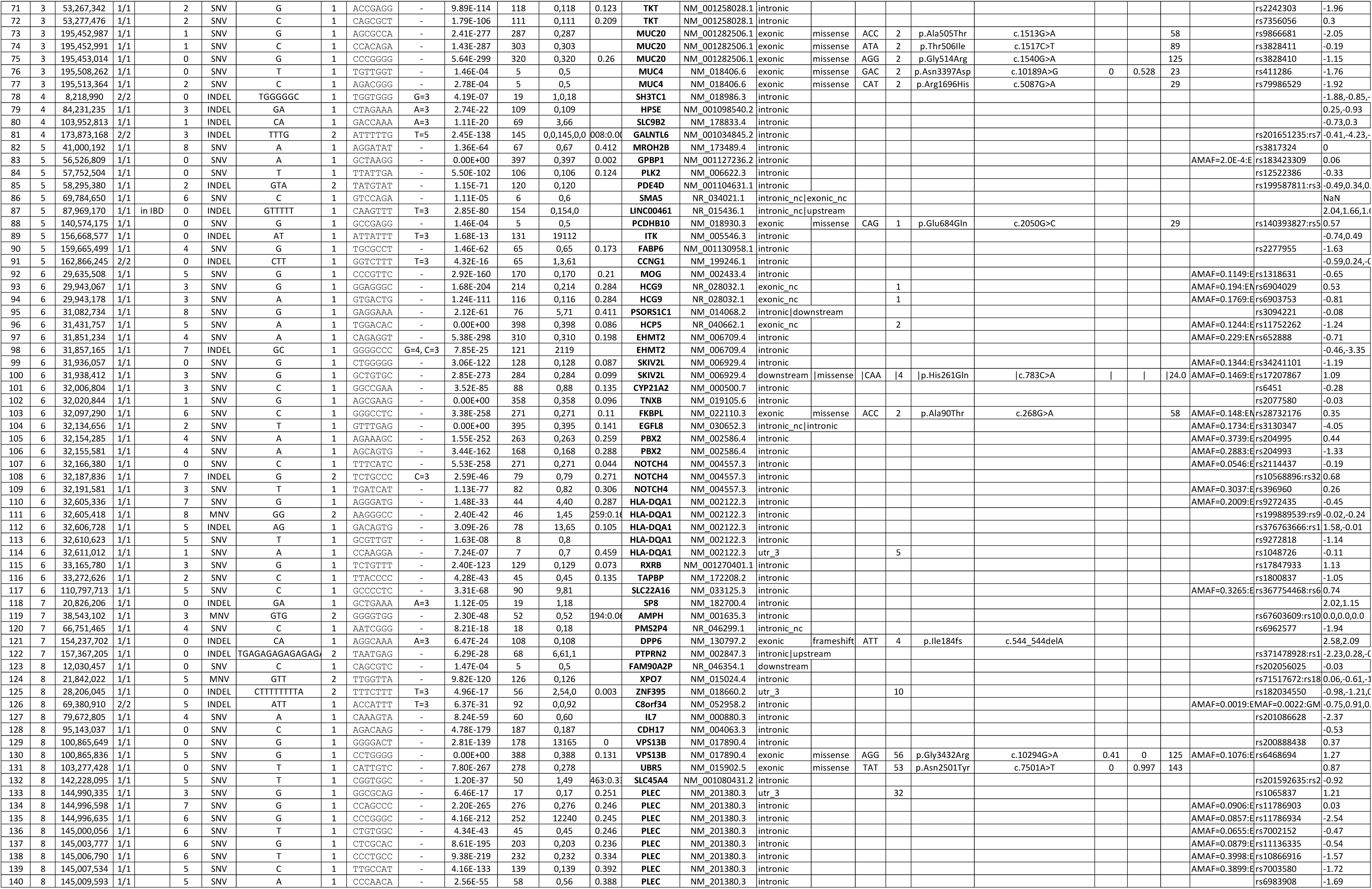
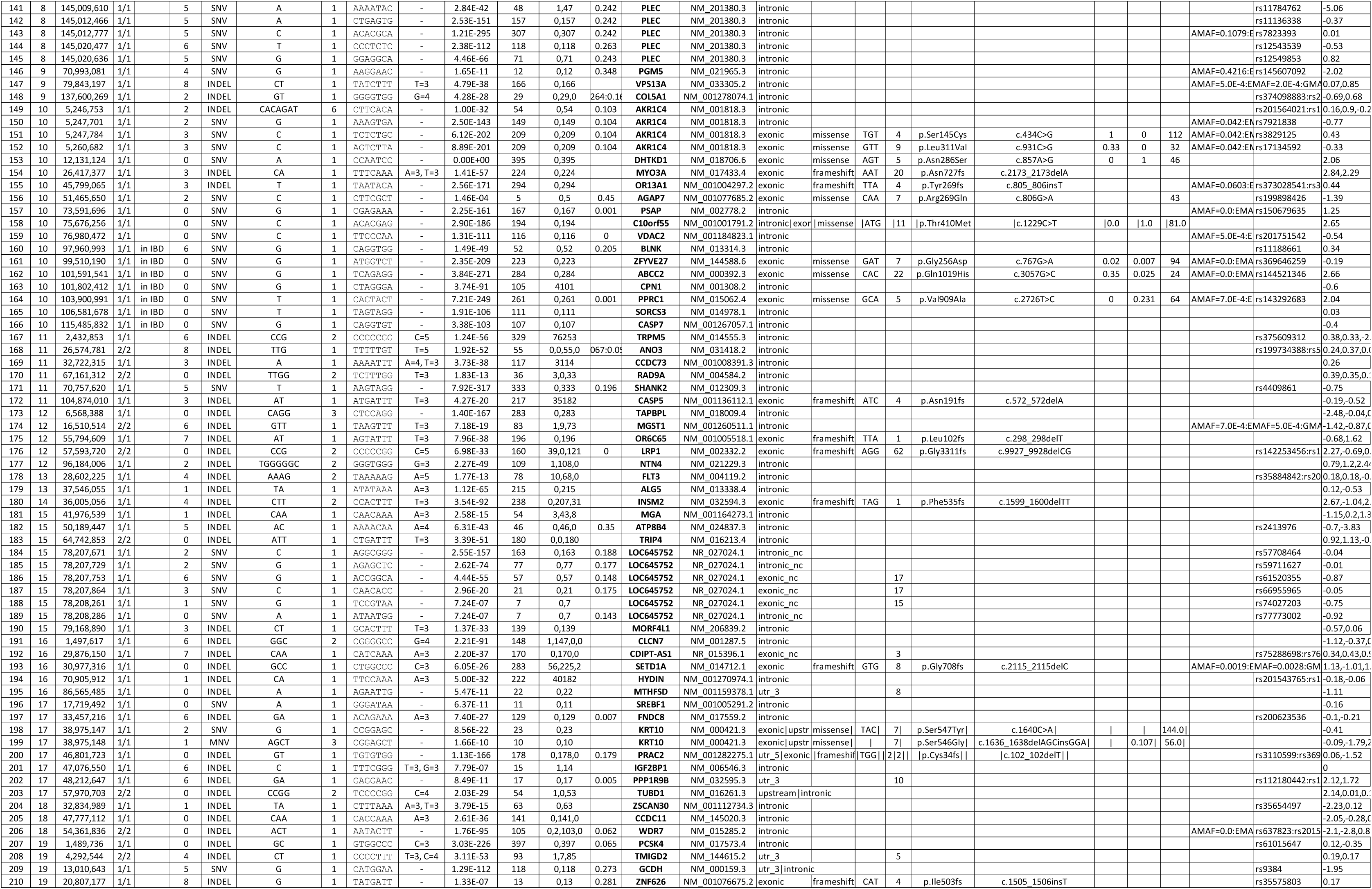
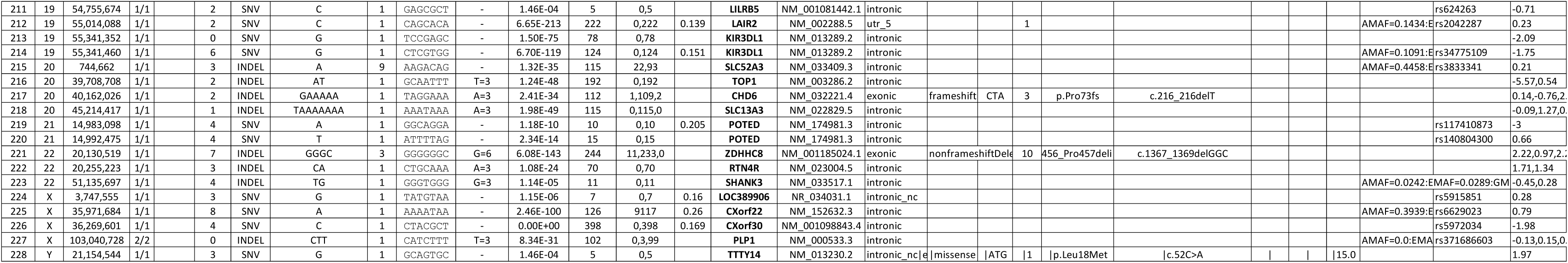
Whole exome sequencing data of patient II.1, with multi-step variant filtering.

*IVNS1ABP* encodes the Influenza Virus NS1A-Binding Protein IVNS1ABP (Q9Y6Y0), also known as NS1-BP or KLHL39, a member of the BTB-Kelch family. Proteins in this family typically contain an N-terminal BTB domain and a C-terminal Kelch domain separated by an intervening linker region known as the BACK domain^32–34^. X-ray crystal structures have been determined for the individual BTB and Kelch domains of IVNS1ABP^35,36^, but not for the full-length protein. The structural models of IVNS1ABP predicted by AlphaFold3^37^ suggested a truncated BACK domain with only its 3-box motif which comprises the first two α-helices. Intriguingly, the remaining region is replaced by a ubiquitin-like β-grasp fold and two intrinsically disordered regions (IDR1 and IDR2), a likely unique feature for this family (Figure 1E). Phenylalanine 253 is located in IDR1, which forms a 51-residue disordered loop within the ubiquitin-like β-grasp domain (Figures 1E and 1F). IDRs often form sites of protein-protein interaction and can undergo disorder-to-order transitions upon complexing with their partners. Mutations in many proteins with IDRs have been implicated in human diseases, such as neurodegeneration^38^. Importantly, Phe253 is conserved across IVNS1ABP orthologues, suggesting that its alteration may disrupt IVNS1ABP-protein interactions.

To examine the consequence of the IVNS1ABP missense variant p.F253C on the binding to one of its direct interacting partners KLHL20^39^, overexpression of IVNS1ABP full-length and KLHL20 Kelch domain in HEK293T cells followed by co-immunoprecipitation was carried out. Unlike wildtype (WT) HA-tagged IVNS1ABP^WT^, the mutant IVNS1ABP^F253C^ could no longer be pulled down by Flag-tagged KLHL20, suggesting that the p.F253C variant may introduce a conformational change in its native folding affecting KLHL20 engagement (Figure 1G). Notably, AlphaFold3 predicted that IVNS1ABP Phe253 forms hydrophobic interactions to KLHL20 in a similar manner as DAPK1 Leu1339 binds to KLHL20^40^ (Figure 1H and S1D). In agreement, previous immunoprecipitation experiments mapped IVNS1ABP residues 234-350, encompassing all of IDR1 and IDR2, as an essential region for many of the native interactions of IVNS1ABP^35^.

Altogether, these results indicate that the conserved region around the disease mutation site is required for IVNS1ABP interaction with its targets, supporting the deleterious effect of the p.F253C variant on IVNS1ABP function.

Past phylogenetic analyses performed on 38 BTB-Kelch family proteins^41^ showed that IVNS1ABP is a close paralogue of the E3 ligase adaptor gigaxonin. The genetic inactivation of gigaxonin (also known as KLHL16, MIM605379) is responsible for Giant Axonal Neuropathy-1 (GAN1, MIM256850), a severe neurodegenerative disease^42^. The comparison of GAN1’s symptomatology with the clinical presentations of this novel syndrome revealed a strong overlap, including a similar progressive peripheral neuropathy leading to muscle weakness, hypotonia and steppage gait. Skin hyperpigmentation, scoliosis, long fingers, club feet, *pes cavus*, distal sensory impairment, dysarthria, reflex dysfunction, mental retardation were all shared between the two diseases (Table 1, Figures 1B and S1D). One of the distinguishing but related phenotypes is that GAN1 patients often present with kinky red hair whereas the three affected patients herein displayed premature hair whitening. The clinical similarities and protein sequence homology between IVNS1ABP and gigaxonin suggested that both proteins may fulfill similar functions by maintaining cellular proteostasis.

### IVNS1ABP^F253C^ induces protein aggregation in patients’ cells

To examine the pathogenicity of this private IVNS1ABP^F253C^ variant, primary fibroblasts isolated from skin biopsies of 2 affected patients (II.1, II.4), a healthy ‘non-carrier’ (II.2) and heterozygous (I.2) family members were reprogrammed into induced pluripotent stem cells (iPSCs). These iPSC lines were then differentiated into disease-relevant cell lineages including neural progenitor cells (NPCs) and post-mitotic neurons (Figure 2A). Using an aggresome detection assay, patients’ primary fibroblasts (IVNS1ABP^F253C/F253C^) showed significant increase of protein aggregates relative to WT cells (IVNS1ABP^WT/WT^) (Figure 2B). This was also observed in the patient’s iPSCs and iPSC-derived neurons (Figures 2C, 2D and S2A). These results suggest that IVNS1ABP^F253C^ prevents proper degradation of endogenous proteins leading to their aggregation across many cell types. To assess the overall cellular proteostasis, we next examined autophagy lysosomal activity which drives the degradation and recycling of cellular wastes. Immunostaining of the lysosome-associated membrane proteins 1 and 2 (LAMP1/2) showed significant enlargement of the lysosomes in each patient cell type studied (Figures 2E and S2B). Increased level of P62, a classical marker of aggrephagy^43^, was also observed by western blot in IVNS1ABP^F253C/F253C^ compared to IVNS1ABP^WT/WT^ and IVNS1ABP^WT/F253C^ NPCs (Figure 2F). These results indicate that maintenance of cellular proteostasis in patients’ cells is significantly altered, causing protein aggregation and overactivation of the autophagy lysosomal pathway.

**Figure 2.**
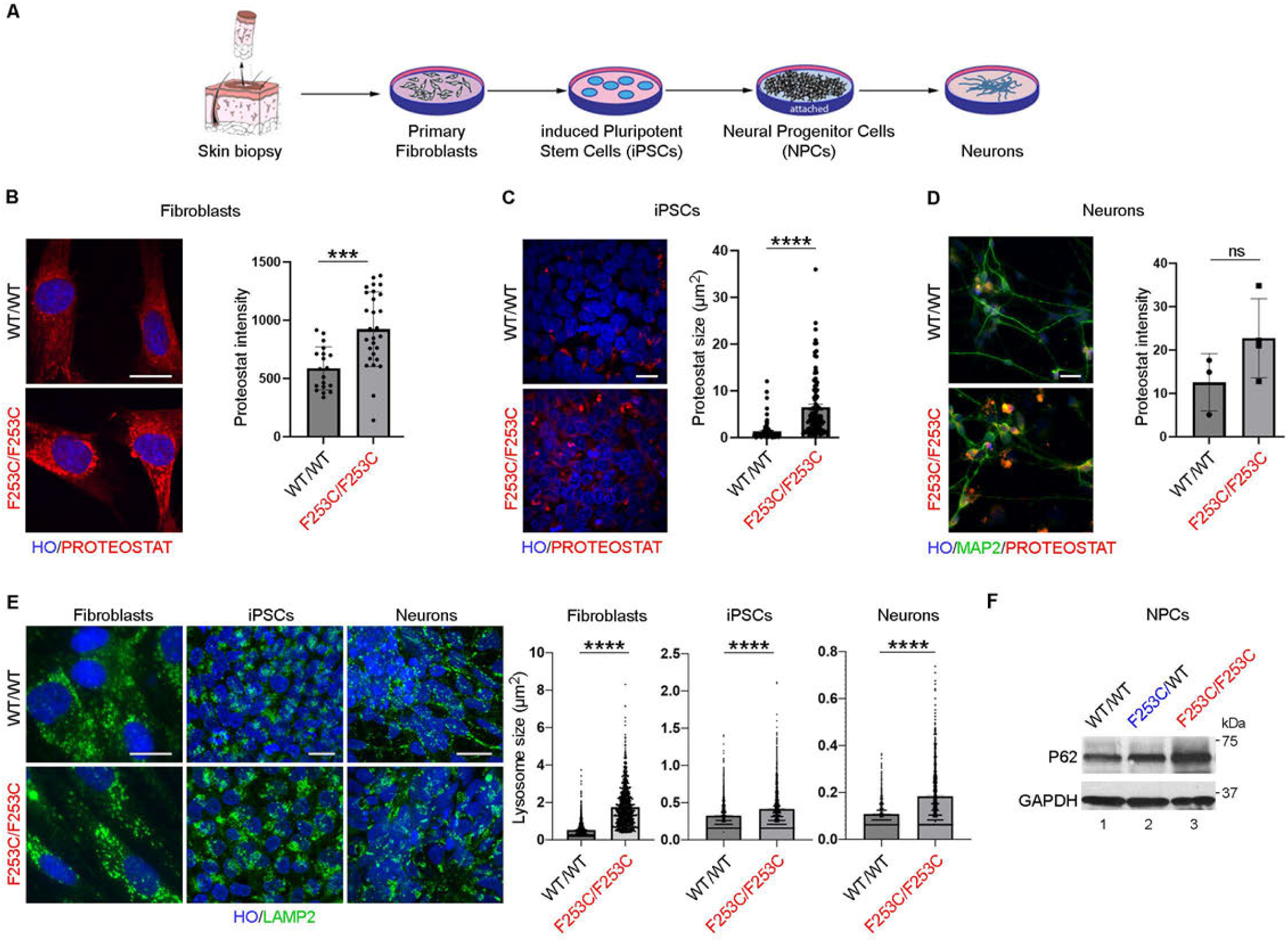
Altered IVNS1ABP induces protein aggregation in patients’ primary cells **(A)** Schematic representation of the different cells generated from punch skin biopsy of patients and controls, from primary fibroblasts, to induced pluripotent stem cells (iPSCs), to neural progenitor cells (NPCs), to differentiated neurons. **(B)** Proteostat® staining and quantification show significant increase of protein aggregates in patient’s primary fibroblasts (II.4) (n=31) compared to unaffected non-carrier cells (II.2) (n=19). Scale bar = 50 µm. Error bars indicate mean ± SEM. Unpaired t test, *** p < 0.001. **(C)** Proteostat® staining and quantification show significant increase of protein aggregates in patient’s iPSCs (II.1) compared to unaffected non-carrier iPSCs (II.2) (each dot represents one protein aggregate. Data were collected from n≥3 independent cultures). Scale bar = 20 µm. Error bars indicate mean ± SEM. Unpaired t test, **** p < 0.0001. See also Figure S2A. **(D)** Proteostat® staining and quantification show increase of protein aggregates in patient’s iPSC- derived neurons (II.1) compared to unaffected non-carrier cells (F.II.2) (each dot represents one culture condition. Data were collected from n=3 independent cultures). iPSC-derived neurons expressed Microtubule-associated protein 2 (MAP2). Scale bar = 50 µm. Error bars indicate mean ± SEM. Unpaired t test, ns: non-significant. **(E)** Immunostaining and quantification of lysosome-associated membrane protein 2 (LAMP2) show significant increase of the lysosomal network in patient’s primary fibroblasts (II.4), iPSCs (II.1) and patient’s iPSC-derived neurons (II.1) compared to unaffected cells (II.2) (One dot represents one lysosome, data were collected from n≥3 independent cultures in each group). Scale bar = 20 µm. Error bars indicate mean ± SEM. Unpaired t test, **** p < 0.0001. See also Figure S2B. **(F)** Western blot of patient’s NPCs (II.4) treated with MG132 shows increased P62 (autophagy marker) compared to unaffected family members (I.1) and (II.2). GAPDH was used as a loading control.

### Loss of IVNS1ABP, or gigaxonin, result in a similar accumulation of ubiquitinated substrates

IVNS1ABP belongs to the BTB-Kelch protein family, members of which serve as adapters for the ubiquitination of protein substrates by multi-subunit E3-ubiquitin ligase complexes. Based on IVNS1ABP’s homology to gigaxonin, we next examined its role in the ubiquitination of protein substrates^33,39,44,45^. Upon cell starvation and Epoxomicin-mediated inhibition of the ubiquitin proteasome pathway (UPP), we found that, compared to WT NPCs, mutant IVNS1ABP^F253C/F253C^ cells displayed evidence of accumulation of ubiquitinated proteins (Figure 3A). To obtain a quantitative measure of these differences and identify possible substrates of IVNS1ABP, we employed targeted mass spectrometry following ubiquitin-mediated enrichment from whole cell extracts (Figure 3B). Significant differences in the total amount of ubiquitinated proteins from the two homozygous patients compared to WT or heterozygous NPCs could be readily quantified (Figures 3C). Remarkably, four of the top 10 differentially ubiquitinated proteins in patients’ NPCs are established substrates of gigaxonin, including Vimentin^46–49^, Tubulin-binding cofactor B^50^,

**Figure 3.**
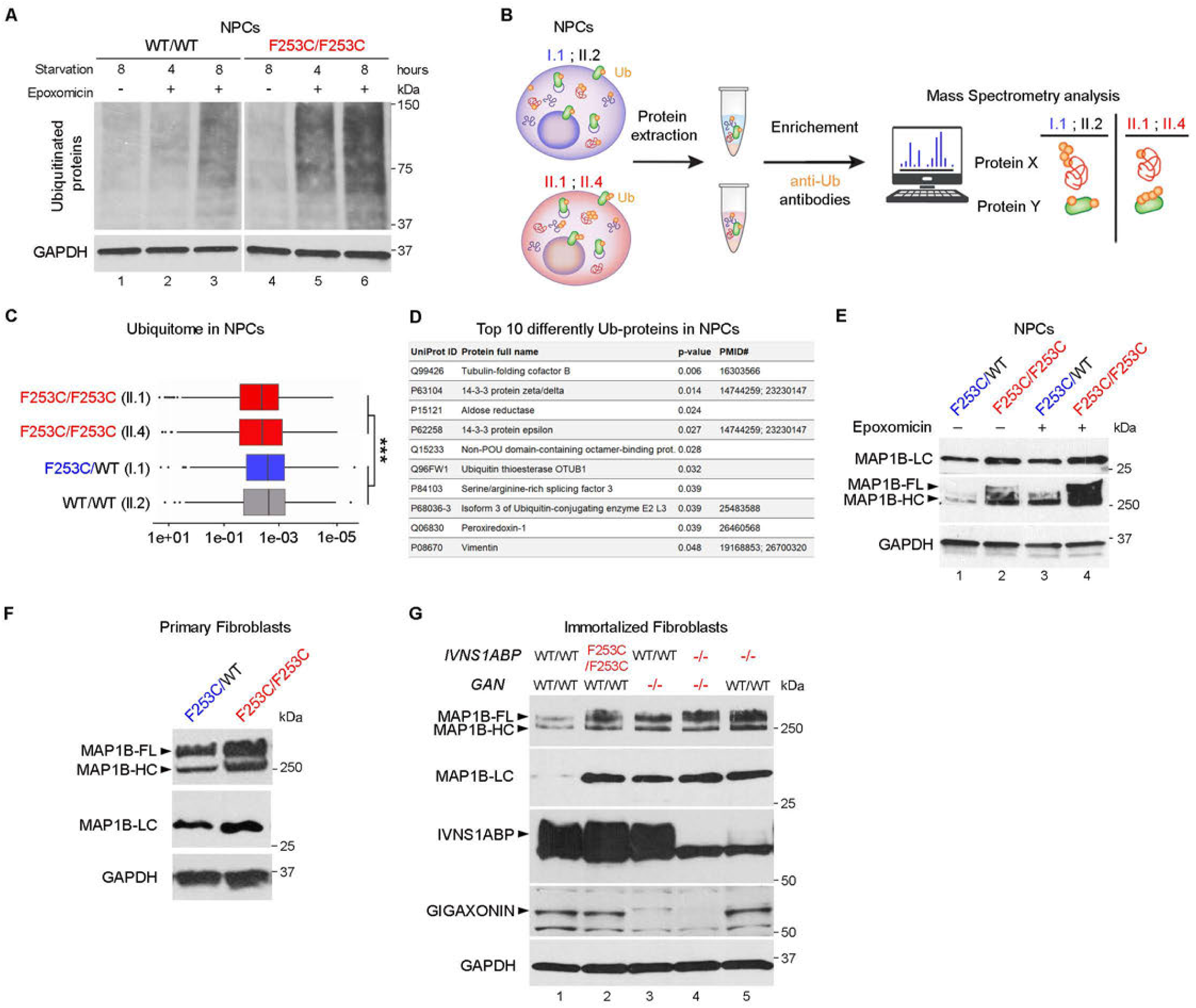
Protein ubiquitination is altered in IVNS1ABP mutant cells **(A)** Western blot of patient’s starved NPCs (II.4), in presence or absence of Epoxomicin, shows increased ubiquitinated proteins compared to unaffected control (II.2). GAPDH was used as a loading control. **(B)** Schematic representation of mass spectrometry workflow to quantify ubiquitinated proteins in whole cell lysates. **(C)** Quantification by mass spectrometry of whole-NPCs lysates from two patients (II.1 and II.4) shows significant increase of ubiquitinated peptides (normalized to total proteins) compared to two healthy controls (I.1 and II.2). Y-axis represents log10 abundances of normalized ubiquitinated peptides. *** p-value<0.001. **(D)** List of the top 10 differentially ubiquitinated proteins in two patients’ NPC extracts (II.1 and II.4) compared to two controls (I.1 and II.2). Five of these are known to be direct substrates of gigaxonin or gigaxonin’s targets. Proteins were ranked by the p-values in ascending order. See also Table S1 and Figure S3. **(E)** Western blot of patient’s NPCs (F253C/F253C, II.4) shows increased endogenous MAP1B compared to whole-cell lysates of unaffected control (WT/WT, II.2), with stronger differences when cells were treated with Epoxomicin. MAP1B LC, Light Chain; FL, Full Length; HC, Heavy Chain. GAPDH serves as loading control. **(F)** Western blot of patient’s primary fibroblasts (F253C/F253C, II.1) also shows increased endogenous MAP1B compared to whole-cell lysates of unaffected control (WT/WT, II.2). **(G)** Western blot of endogenous IVNS1ABP, MAP1B and gigaxonin in immortalized fibroblasts of unaffected control II.2 (WT/WT, lane 1), *IVNS1ABP*-mutated patient II.1 (F253C/F253C, lane 2), *GAN*-null patient (-/-, lane 3), CRISPR-Cas9-induced *IVNS1ABP* knockout (-/-) in *GAN*-null patient (-/-, lane 4) and CRISPR-Cas9-induced *IVNS1ABP* knockout (-/-) in unaffected control II.2 (WT/WT, lane 5). All cell lines show increased MAP1B compared to unaffected WT/WT control II.2. MAP1B LC, Light Chain; FL, Full Length; HC, Heavy Chain. GAPDH was used as a loading control.

Peroxiredoxin-1^46^ and 14-3-3 proteins^51^ (Figure 3D, Table S1 and Figure S3). Other hits included the ubiquitin-conjugating enzyme E2 L3 (UBE2L3) which was shown to bind MAP1B, a prototypical substrate of gigaxonin^52^ (Figure 3D). This prompted us to evaluate the levels of MAP1B in patients’ NPCs by western blot. We observed that, with or without UPP inhibition by Epoxomicin, MAP1B full length or its cleaved light and heavy chains, were noticeably increased in patient cells relative to heterozygous IVNS1ABP^F253C/WT^ cells (Figure 3E). The accumulation of MAP1B being a tell-tale sign of Gixagonin deficiency in GAN1 patient-derived cells, we sought to see if this was also true in patient’s primary fibroblasts. Compared to heterozygous fibroblasts, endogenous MAP1B polyprotein and subunits were robustly increased in IVNS1ABP^F253C/F253C^ cells (Figure 3F). These results support the notion that IVNS1ABP and gigaxonin earmark similar substrates for degradation, explaining why their genetic inactivation results in a similar set of neurodegenerative phenotypes.

To address whether Gixagonin and IVNS1ABP are functionally redundant, we sought to characterize single and double knockout fibroblasts for these two paralogous genes. First we derived fibroblasts from a GAN1 patient with a validated null homozygous allele which deletes the entirety of the *GAN* locus (breakpoints [GRCh37]16q23.2 [81262391_81445961]). The *GAN*-null patient showed similar musculoskeletal defects, progressive and severed neuropathy as did the three IVNS1ABP-deficient patients (Figure S1D and Table 1). By CRISPR-Cas9, we then knocked out *IVNS1ABP* in WT or *GAN*-null cells. These engineered and immortalized fibroblasts were confirmed to be single or double KO for *IVNS1ABP* and/or *GAN* by genomic sequencing (data not shown) and by western blotting (Figure 3G). Using endogenous MAP1B levels as a proxy for IVNS1ABP and gigaxonin activity, we observed that IVNS1ABP^F253C/F253C^ (lane 2) and IVNS1ABP^KO/KO^ (lane 5) cells equally result in the accumulation of MAP1B, indicating that this recessive missense allele behaves as KO allele. As expected, the loss of gigaxonin (lane 3) resulted in accrued MAP1B levels, which was comparable to that of IVNS1ABP KO cells, but was not further enhanced in *GAN/IVNS1ABP* double KO cells (lane 4) (Figure 3G). These unambiguous KO alleles and pairwise isogenic cell lines proved to be robust readouts for addressing the complementarity of gigaxonin and IVNS1ABP. We find that both E3 ligase adapter proteins are needed for the degradation of common cytoskeletal substrates such as MAP1B, the accumulation of which may lead to neurotoxicity.

### Axonal damage is rescued in genetically-corrected IVNS1ABP motor neurons

To further prove the pathogenicity of this private IVNS1ABP variant and mitigate the effect of possible genetic modifiers, isogenic lines were created using CRISPR-Cas9. To this end, the homozygous mutation (c.758T>G; p.F253C) in patient’s iPSCs (II.1) was genetically and bi- allelically corrected into its WT genotype (c.758T/T)^53^ (Figure 4A). Patient-derived and their isogenic WT iPSCs (WT^corr.^/WT^corr.^) efficiently differentiated into motor neuron (MN) progenitors, expressing MN markers such as OLIG2 and MNX1^54^ (Figure S4). After 4 weeks of culture, these paired isogenic MN progenitors differentiated into mature MNs expressing choline acetyltransferase (ChAT) (Figure 4B). Less abundant and enlarged neurite bundles extending from the MN spheres marked by MAP2 were observed in mutant IVNS1ABP^F253C/F253C^ cells compared to isogenetically-corrected WT ones (Figure 4B). As previously observed in non-isogenic fibroblasts, iPSCs and neurons (Figure 2E), enlarged lysosomes in IVNS1ABP^F253C/F253C^ MNs compared to isogenic WT were documented (Figure 4C). The use of a lysotracker and live imaging revealed slower intra-cellular movement of lysosomes which was rescued in IVNS1ABP^Corr./Corr.^ MNs (Figure 4D). Immunostaining of neural filaments (NF) using phosphorylated p-NF, an established marker of axonal damage^55^, showed abnormal p-NF accumulation especially in the distal ends of mutated axons, suggesting potential axon transport deficit (Figure 4E). Western blot analysis confirmed a noticeable increase of p-NF triple proteins (heavy, medium and light chains) in patient’s MNs compared to WT corrected ones (Figure 4F).

**Figure 4.**
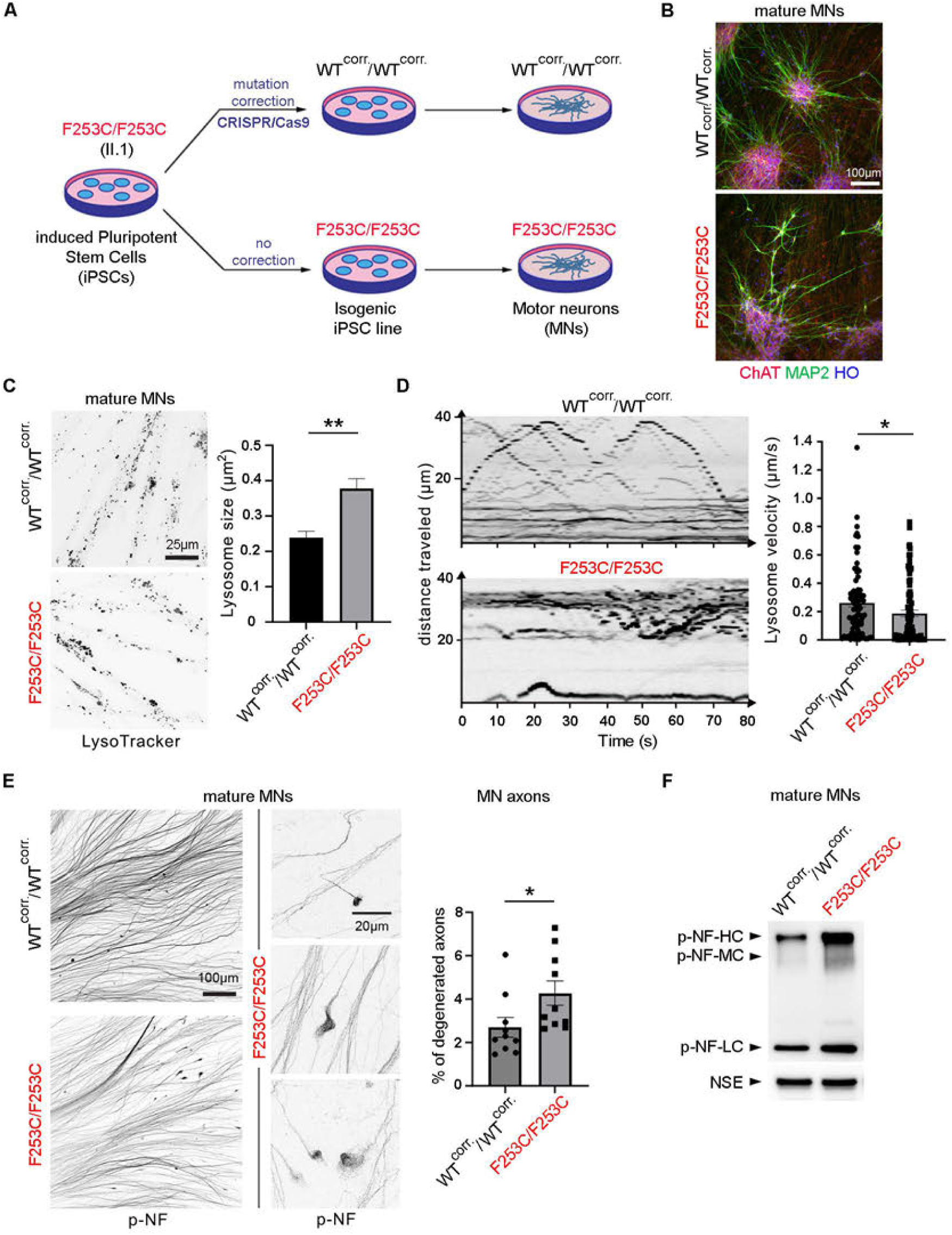
*IVNS1ABP* mutation leads to human motor neuron degeneration **(A)** Mutation F253C/F253C in patient’s iPSCs (II.1) was corrected using CRISPR-Cas9, creating isogenic control iPSCs (WT^corr.^/WT^corr.^). Spinal cord motor neurons (MNs) were differentiated from both isogenic iPSCs. **(B)** Immunostaining of ChAT and MAP2 shows that isogenic motor neuron progenitors differentiated further into mature MNs. **(C)** Lysotracker representative image and quantification show significant increase of the lysosome size in patient’s iPSC-derived MNs (F253C/F253C) compared to its isogenic wildtype cell line (WT^corr.^/WT^corr.^). Error bars indicate mean ± SEM. Unpaired t test, ** p < 0.01 (n=5 culture conditions). **(D)** Live imaging of lysotracker dye in iPSC-derived MNs (4-week-old) shows impaired intracellular lysosome movement over time in patient’s MNs (n=87) compared to its isogenic wildtype MNs (n=80). Each dot represents one organelle labeled by lysotracker. Data was collected across three independent cultures. **(E)** Immunostaining with phospho-neurofilament antibodies (p-NF) shows that mutant F253C/F253C MNs display p-NF accumulation at the distal ends of MNs compared to isogenic WT^corr.^/WT^corr.^ cells. Each data point represents the ratio of damaged axons (labeled with pNF aggregates) in each image field taken from 3 independent MN cultures. Error bars indicate mean ± SEM. Unpaired t test, * p < 0.05. **(F)** Western blot of patient MNs show increased p-NF compared to its isogenic control cells. p- NF-H, heavy chain; p-NF-M, medium chain; p-NF-L, light chain. NSE, a neuron-specific enolase, is used as a loading control.

p-NF is a pathological hallmark for axonal damage in multiple neurodegenerative diseases, such as amyotrophic lateral sclerosis, Alzheimer’s disease and GAN1^56,57^. Altogether these *in vitro* experiments using isogenic pairs of iPSC-derived MNs confirmed the deleterious effect of p.F253C, which we speculate may be responsible for motor neuron degeneration in the three affected patients.

### *Ivns1abpa/b* KO zebrafish show defective motor neuron development and locomotion

To better understand the physiological role of *Ivns1abp*, we next sought to create an animal model of the disease. We favored zebrafish over mice, following the observation that *gan* KO fish more faithfully recapitulated the severity of human disease than did mouse models of GAN1^58–61^. In the zebrafish genome, *Ivns1abp* has been duplicated into *ivns1abpa* (GenBank NM_199279) and *ivns1abpb* (GenBank NM_201483), which are situated on chromosome 20 and 2 respectively. The spatio-temporal expression of both genes was analyzed by RT q-PCR and wholemount *in situ* hybridization. *Ivns1abpb* was highly expressed in zygotes indicating maternal deposition in the egg, while *ivns1abpa* was expressed from 3-somites stage onwards (Figures S5A and S5B). While phylogeny and synteny suggest that *ivns1abpa* is closer to the human *IVNS1ABP* locus (Figure S5C), we knocked out both genes using CRISPR-Cas9. We identified, selected, and outbred independent germline frameshift alleles for each gene: *ivns1abpa^7bpdel^:* c.195delGGAACCT; p.(Glu66Metfs*5) and *ivns1abpb^2bpdel-1bpins^:* c.205delGGinsT; p.(Gly69Leufs*27) (Figure S6A). Each mutated fish line was bred to homozygosity and intercrossed allowing to generate double maternal zygotic *ivns1abpa/b* null fish (referred to as MZ dKO) (Figure S6B). QPCR and western blot analyses of MZ dKO embryos indicated that both transcripts were targeted for nonsense mediated decay, resulting in *ivns1abpa/b* protein-null fish (Figures 5A, 5B and S6C). MZ dKO fish did not display overt anatomical differences during early embryogenesis but exhibited a shortened lifespan with approximately 45% dying within the first month post fertilization. If bred in isolation, a few MZ dKO escapees could be grown up to one year of age (Figure 5C). Compared to WT, MZ dKO fish had a propensity to swim less and remained at the bottom of tanks. Based on the peripheral neuropathy diagnosed in the three IVNS1ABP^F253C/F253C^ patients and the cellular defects observed in patient-derived MNs, we next assessed the development of primary MNs in zebrafish larvae. During the first day of development, neuromuscular connections are limited to three primary MNs per hemisegment, including caudal primary (CaP) MN which is the only one extending an axon toward skeletal muscles, leading to spontaneous motor activity^62,63^. We therefore investigated the morphology of CaP MNs at 32 hours post fertilization (hpf) using an anti-Synaptotagmin 2 antibody^64^. Microscopic observation of trunk hemisegments between somites 2-7 revealed a significant increase of poorly fasciculated CaP MNs in MZ dKO (90%) compared to control (28%). Aberrant CaP axon pathfinding and ectopic branching in MZ dKO (35%) compared to control (5%) was also readily documented and quantified (Figure 5D). These abnormal primary MN phenotypes evoke those seen in *gan* knockout fish^59^, indicating that *ivns1abp* may equally be required for proper axonal development of MNs.

**Figure 5.**
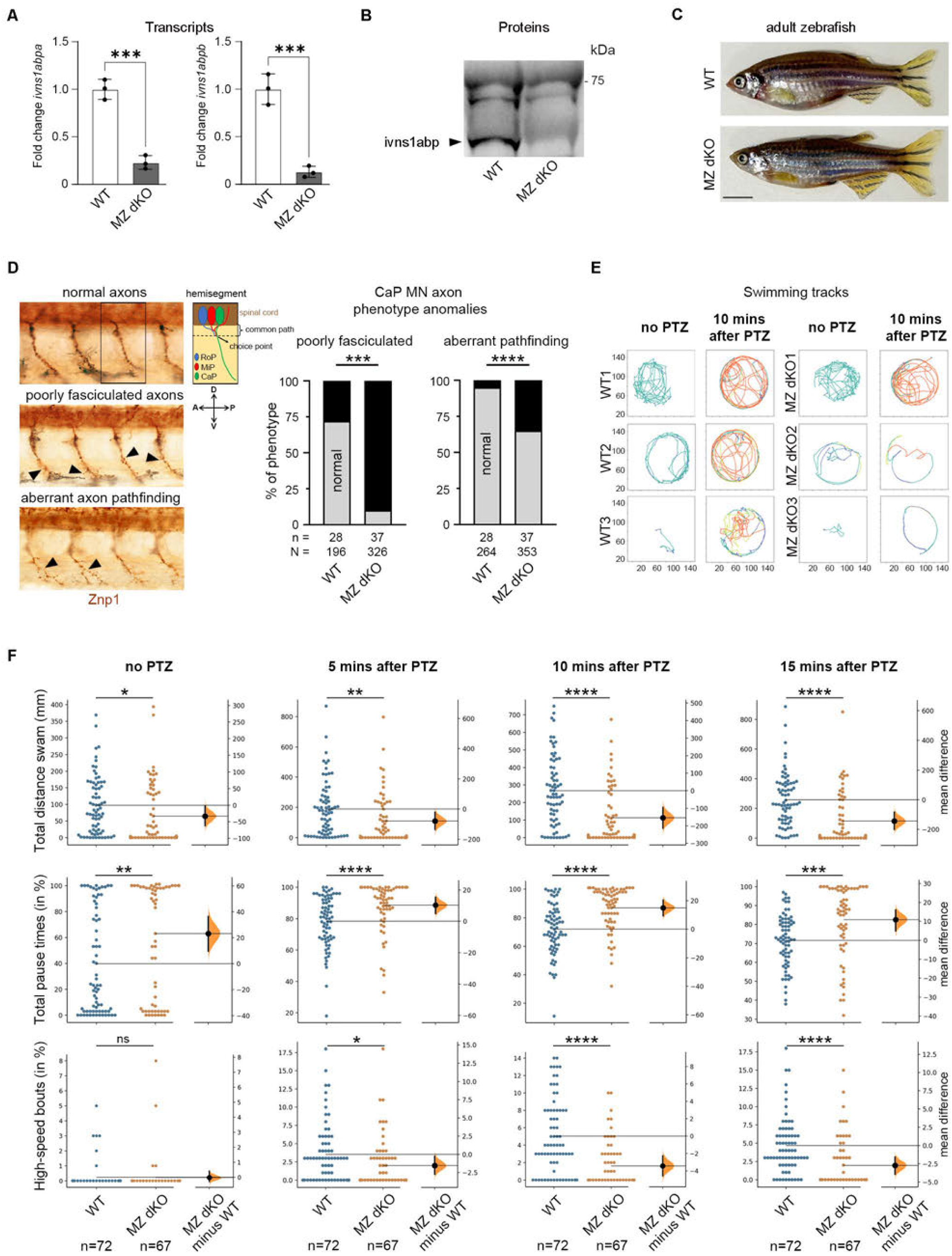
Maternal zygotic double knockout of *ivns1abpa and ivns1abpb* leads to defective motor neuron development in zebrafish **(A)** qRT-PCR of maternal zygotic double knockout (MZ dKO) embryos at 24 hours post-fertilization (hpf) shows depletion of both *ivns1abpa* and *ivns1abpb* transcripts. See also Figure S6A. Unpaired t test, *** p < 0.001, n=3 biological samples, with at least 20 embryos for each biological sample. **(B)** Western blot of MZ dKO embryos at 1-day post-fertilization (dpf) shows absence of ivns1abp proteins. See also Figure S6B. **(C)** Representative image of 1-year-old WT and MZ dKO female fish shows no obvious phenotypic difference. Scale bar, 1000 µm. **(D)** Representative confocal z-stack images of 4 hemisegments of 31 hpf WT and MZ dKO embryos immunostained with Znp1, as well as a schematic representation of the rostral, middle and caudal primary MNs (RoP, MiP and CaP respectively) with their axon trajectories in each hemisegment (A: anterior, P: posterior, D: dorsal, V: ventral). Compared to normal CaP motor neuron axons, MZ dKO embryos show higher % of poorly fasciculated ventral axons (*** p < 0.001) and aberrant pathfinding (**** p < 0.0001), with mild to severe projections that branched out. **(E)** Exemplary swimming paths extracted from 2-minute videos of three WT and three MZ dKO 7 dpf larvae, with each larvae in individual well of a 24-well plate. WT larvae exhibited higher swimming distance and speed compared to MZ dKO, especially after 10 minutes exposure of 5 mM PTZ. Colors represent different velocity ranges, with blue: low (< 8mm/sec), yellow: intermediate (8-16 mm/sec), and orange: high (> 16mm/sec). x and y Axis represent pixels. **(F)** Compared to WT (n=72), 7 dpf MZ dKO (n=67) larvae show significant differences in locomotor behaviors, including shorter distance swam, higher total pause time between swim bouts, and lower high-speed bouts where velocity was at least 16 mm/s. Larvae were challenged with 5 mM of Pentylenetetrazol (PTZ) for 5, 10, and 15 minutes (mins). In Gardener-Altman and Cummings estimation plots, the mean difference between the two groups (black dot) and 95% confidence intervals (black bar) are plotted on the secondary axis. The orange shaded area in orange shows the 5000 bootstrapped confidence intervals. Two-sided permutation t-test for p-values are represented as * p < 0.05, ** p < 0.01, *** p < 0.001, **** p < 0.0001. See also Figure S6D.

Since MNs extend their axons toward skeletal muscles, leading to spontaneous motor activity, we next assessed whether these developmental anomalies could impact fish motility. At 7 days post fertilization (dpf), MZ dKO larvae (n=67) showed weaker locomotor capabilities, characterized by significantly lower distance swam and longer pause times compared to WT larvae (n=72) (Figures 5E, 5F and S6D). These differences were even more pronounced at 5, 10 and 15 minutes post Pentylenetetrazol (PTZ) treatments, suggesting that at baseline and upon sensitization, MZ dKO fish exhibited impaired locomotion and overall swimming behaviors. These results are consistent with previous findings where defective axon pathfinding and axon over-branching in *ccdc80-l1* or *gan* knocked-down zebrafish embryos led to motility issues^64^. Altogether, these *in vivo* results obtained in a surrogate animal model revealed that the physiological function of ivns1abp towards CaP MN development is conserved between mammals and fish. The complete inactivation of both *ivns1abpa/b* genes in zebrafish complements and supports the notion that the identified missense variant in humans is behaving as a typical loss-of-function allele.

## Discussion

Here we identify IVNS1ABP as a critical regulator of cellular proteostasis. A private germline homozygous variant in *IVNS1ABP* in three affected patients was found to segregate with a new syndrome, consisting of progredient peripheral neuropathy, skin dyschromatosis and premature hair whitening. The complete genetic ablation of both *ivns1abp* genes in zebrafish phenocopies the human disease, resulting in defective locomotion which could be traced back to defective establishment of primary motor neurons. On a cellular level, the alteration of p.F253C caused significant accumulation of protein aggregates in human fibroblasts, iPSCs and NPCs. Remarkably the isogenic correction of the identified mutation in iPSC-derived MNs was sufficient to rescue these cellular vulnerabilities. Targeted proteomic analysis revealed a number of improperly ubiquitinated proteins in IVNS1ABP-deficient cells, including known targets of gigaxonin, which we propose works in concert with IVNS1ABP to prevent the accumulation of protein aggregates.

Recessive loss-of-function variants in *GAN* cause Giant axonal neuropathy-1 (GAN1), a disease that displays strikingly similar clinical features and functions as this IVNS1ABP-related syndrome^42,65,66^. In humans, sporadic heterozygous mutations in *IVNS1ABP* have been associated with primary immunodeficiency^67^. No recessive disease causative mutation in *IVNS1ABP* has been described before. Interestingly, it is worth noting that most of the KLHL proteins are expressed in the nervous system and play a role in neuronal structure and function^44^. Our recent findings of patients lacking KBTBD2 with a neurodevelopmental disorder further support this notion^68^. This suggests that IVNS1ABP, like gigaxonin and other KLHL proteins, may be central for neuronal development, function and homeostasis.

### Are IVNS1ABP and gigaxonin functionally redundant?

The protein structure homology, the overlapping clinical presentations and shared protein substrates, point to a hitherto un-appreciated functional relatedness between IVNS1ABP and gigaxonin.

Our ubiquitome analysis revealed Tubulin-folding cofactor B (TBCB) as the most differently ubiquitinated in patients’ cells compared to healthy controls. TBCB was shown to interact with gigaxonin for its targeted degradation via the UPP^50^. TBCB is well known to play a role in neuronal growth cones, controlling microtubule dynamics and plasticity^69^. Whereas TBCB knockdown promotes axon growth, TBCB overexpression leads to microtubule depolymerization and axonal damage, which are witnessed in GAN1 patients^70^. Vimentin, Peroxiredoxin-1 (PRDX1) and 14-3- 3 proteins are additional IVNS1ABP targets that overlap with previously published gigaxonin pull-down experiments^46,51^. For instance, Vimentin accumulation in gigaxonin-deficient patients’ fibroblasts^47^, a tell-tale sign of GAN1, was readily observed in IVNS1ABP patient’s cells as well. It is worth mentioning that the aggregation of mutant Vimentin in a patient carrying a heterozygous p.(Leu387Pro) variant is also associated with a progeroid syndrome with motor and sensitive peripheral neuropathy^71^. This suggests that the accumulation of Vimentin might be an important driver of the pathogenesis of IVNS1ABP deficiency. Similarly, the dysregulation of the antioxidant PRDX1 enzyme, known to control motor neuron differentiation^72^, and 14-3-3 proteins which interact with neurofilament protein-L and regulate dynamic assembly of neurofilaments^73^ were further evidence that IVNS1ABP and gigaxonin converge on a similar set of substrates.

MAP1B is another well-established substrate of gigaxonin. Mutations in the KELCH domain of gigaxonin were shown to disrupt its interaction with MAP1B, leading to its accumulation and abnormal elongation and guidance of axons^74^. Upon synthesis, MAP1B full length (FL) is cleaved in MAP1B heavy chain (MAP1B-HC) which possesses a microtubule binding site (MTB) at its N-terminus, and MAP1B-LC which contains an actin binding site in its C-terminus^74,75^. Although MAP1B was not part of the top 10 targets in our ubiquitome analysis, the Ubiquitin-conjugating enzyme E2 L3 (UBE2L3), a known binding partner of MAP1B-LC that favors UBE2L3-mediated UPP^52^, significantly accumulated in patients’ NPCs compared to control cells. Accordingly, we found that endogenous MAP1B (FL, HC and LC) protein levels across different cell types significantly accumulated in *IVNS1ABP*-mutated and -null cells, as they did in *GAN*-null patient’s fibroblasts. Our results documenting similar MAP1B and Vimentin accumulation in IVNS1ABP and gigaxonin single knockout fibroblasts argue in favor of both being needed for their effective turnover. However, one may be surprised that double *IVNS1ABP/GAN* knockout cells do not show worsened MAP1B levels than what is seen in single KOs. This indicates that IVNS1ABP alone cannot compensate for the loss of gigaxonin and vice versa. This is supported at the organismal level in humans or zebrafish, where the inactivation of each gene produces a phenotype that is not entirely masked by the remaining active paralogue. To examine the full extent of a possible functional redundancy between IVNS1ABP and gigaxonin, triple KO zebrafish or double KO mice will need to be generated to better appreciate if compensatory mechanisms take place, which would speak to the need of having both active paralogues to enforce tight proteostasis. Notwithstanding, our results indicate IVNS1ABP and gigaxonin are responsible for protein clearance of common substrates, which is reflected in the strong overlapping symptomology seen in both syndromes.

### The moonlighting function of IVNS1ABP

Besides controlling proteostasis, IVNS1ABP seems to play different roles in multiple biological pathways, including spliceosome machinery^35,76,77^, signal transduction^78^, c-Myc transcriptional regulation^79^, and actin stabilization and cytoskeleton organization^80^. Concurrent investigation by Fang *et al.,* (2024) using the same isogenic patient cell lines carrying homozygous WT, p.F253C or KO alleles of IVNS1ABP showed that this E3 ligase is also regulating actin dynamics. For instance, IVNS1ABP^F253C/F253C^-mutated or IVNS1ABP^KO/KO^ cells displayed dysregulation of actin polymerisation, which led to impaired cytokinesis and mitosis, resulting in cell cycle arrest and premature senescence. All in all, our parallel investigations have revealed IVNS1ABP’s function towards maintaining cellular homeostasis by controlling the levels of key cytoskeletal proteins.

### Therapeutic intervention

Owing to the molecular, cellular and clinical signatures shared by *GAN*- and *IVNS1ABP*-deficient patients, we suspect that similar modalities could be employed for therapeutic interventions. Recently a small molecule screen in morpholino-depleted and knockout *gan* zebrafish has identified a few hit compounds^59^. These small molecules belong to the class of muscarinic antagonist and α-adrenergic agonist and proved to be efficacious in stabilizing the neuro- muscular junction and increasing the locomotion of *gan* deficient fish. Future experiments will need to assess whether these molecules can also prove to be effective in rescuing the locomotion in double *ivns1abpa/b* mutant fish, which would lend further credence to the notion that both diseases share a common pathogenesis.

The investigational gene therapy led by the NIH involving the single injection of a modified scAAV9/JeT-GAN virus into the fluid surrounding the spine of GAN1 children is an alternative strategy to restore adequate levels of gigaxonin^81^. The promising observation of a dose-dependent slowing down of motor decline in treated children suggest that similar approaches could be envisaged for IVNS1ABP-deficient patients which may also benefit from an exogenous supply of gigaxonin.

## Data Availability

All data produced in the present work are contained in the manuscript

## Acknowledgments

We are grateful to all members of the Reversade laboratories for support, especially Pui-Mun Wong for technical advice, and Luc Grometto for movie editing. We thank Hane Lee, Barry Merriman, and Ascia Eskin (University of California Los Angeles) for the SNP-genotyping and homozygosity mapping. We thank Christoph Wolfram Winkler for his advice on zebrafish motor neuron study. C.B. is supported by a NMRC Open Fund - Young Individual Research Grant (OF- YIRG/0048/2017). F.Y. is supported by a NMRC Open Fund - Young Individual Research Grant (OF-YIRG22jul-0021) and Duke-NUS-KPFA/2020/0038. S-C.Z. acknowledges the support from Singapore Ministry of Education, MOE2018-T2-2-103 and Singapore Ministry of Health, MOH- 000207 and 000212. A.S.M. acknowledges the support from Ministry of Education MOE- T2EP30220-0020. B.R. is a fellow of the National Research Foundation (NRF, Singapore) and Branco Weiss Foundation (Switzerland) and an EMBO Young Investigator. Z.C acknowledges support from the China Scholarship Council - Nuffield Department of Medicine Scholarship. A.N.B. acknowledges support from Wellcome (106169/ZZ14/Z). This work is supported by the Biomedical Research Council (BMRC), Agency for Science, Technology and Research (A*STAR) core funding and Singapore National Research Foundation grant (NRF-SIS) to RMS. This work was funded by a Strategic Positioning Fund for Genetic Orphan Diseases (SPF2012/005) and an inaugural A*STAR Investigatorship from the Agency for Science, Technology and Research in Singapore to B.R.

## Author contributions

M.S., U.A., M.E.K., A.M., G.T.T. and R.F. made clinical diagnoses and collected clinical data and samples. M.W.Y., A.Y.J.N., S.T., V.B. and M.C. performed SNP genotyping, WES and Sanger sequencing on the family. B.K. molecularly diagnosed the *GAN*-null patient. P.W.C. and G.N. reprogrammed fibroblasts into iPSCs and derived NPCs. C.B. and A.N.A. performed all the experiments and analysis on primary fibroblasts and neural progenitor cells. C.B., A.N.A., N.R.,

N.A.B.M.K and E.S-R performed zebrafish work. A.S.M. and V.C. performed zebrafish locomotion assay and axon staining. F.Y., D.X., Y.S.T, S.M.C and S-C.Z. designed and performed iPSCs derived MN experiments and characterized lysosome and protein aggregation. Z.C. and A.N.B. analyzed KLHL20 binding by co-IP and AlphaFold predictions. R.M.S., K.Y.T., Y.T.L. performed ubiquitination assay and analyzed mass spectrometry data. C.B. and B.R. designed the study and wrote the manuscript with input from all co-authors.

## Competing interest

The authors declare no competing interests.

## **STAR Methods** (format not required for first submission)

### Human study participants

Peripheral blood samples were withdrawn from 8 members of the family to extract genomic DNA (gDNA) using DNeasy Blood & Tissue Kits (Qiagen). Skin biopsies from two patients II.1, II.4, unaffected family members I.1 and II.2 were collected to isolate primary skin cells. Cutaneous fibroblasts were derived from a GAN1 male patient with a recessive deletion 16q23.2 [81262391- 81445961] (GRCh37) which encompasses the entirety of the *GAN* gene. All bio-specimens were obtained after written informed consents were signed from participants or their legal guardians. All human studies were reviewed and approved by the institutional review board of A*STAR (2019-087).

### Homozygosity mapping and exome sequencing of the family

Eight family members (I.1, I.2, II.1-II.6) were genotyped using Illumina Humancore-12v1 BeadChips following manufacturer’s instructions. Call rates were above 99%. Gender and relationships were verified using Illumina BeadStudio. Mapping was performed by searching for shared regions that are homozygous and identical-by-descent (IBD) in the three affected patients using custom programs written in the Mathematica (Wofram Research, Inc.) data analysis software. As previously described, candidate regions were further refined by exclusion of common homozygous segments with any unaffected family members^82^. The confidence criteria to identify IBD blocks were a minimum of 2 cM. We identified four shared candidate loci on chromosomes 1, 5, 10 and 15, totalling 93.7 Mb. Exome sequencing was performed on gDNA from patient II.1. The library was prepared on an Ion OneTouch System and sequenced on an Ion Proton instrument (Life Technologies). Sequence reads were aligned to the human GRCh37/hg19 assembly (UCSC Genome browser). Each variant was annotated with the associated gene, location, protein position, amino acid change, quality-score and coverage. Variants were filtered for common SNPs using the NCBI’s “common and no known medical impacts” database (ftp://ftp.ncbi.nlm.nih.gov/pub/clinvar/vcf_GRCh37/), the Genome Aggregation Consortium (https://gnomad.broadinstitute.org/), and the Exome Sequencing Project (http://evs.gs.washington.edu/EVS/), as well as an in-house database of 666 sequenced individuals, mainly of Middle East origin. Homozygous variants were further filtered based on functional prediction scores including SIFT^83^, PolyPhen-2^84^, PhyloP conservation scores^85^ and Mendelian Clinically Applicable Pathogenicity (M-CAP)^86^. Sanger sequencing using primers flanking the mutation (Forward: 5’-GCTGCTTGATGGGAACCTAC-3’; Reverse 5’- GCTTGTTTTCTCCCAAAAATG-3’) confirmed segregation of the variant with the disease.

### Co-immunoprecipitation

HA-tagged human *IVNS1ABP* wildtype (HA-IVNS1ABP^WT^) and mutant (IVNS1ABP^F253C^) cDNAs were synthesized from mRNA extracted from control and patient fibroblasts using GenScript and cloned into pCS2+ vector. Flag-tagged human KLHL20 Kelch domain M303-W609 (Uniport identifier Q9Y2M5-1) was subcloned into pcDNA3-N-Flag-LIC vector. HEK293T cells were cultured in Dulbecco’s modified Eagle’s medium (DMEM, Gibco/Invitrogen) supplemented with 10% fetal bovine serum (FBS, Sigma Aldrich), 100 U/mL penicillin sodium and 100 µg/mL streptomycin sulfate (Sigma Aldrich) in a humidified incubator at 37°C with 5% CO2. KLHL20 and IVNS1ABP were co-transfected 1:1 into HEK293T cells with a total amount of 5 µg DNA and 15 µg Polyethylenimine (PEI) at 60% cell confluency. Cells were harvested 40 h post transfection and lysed in an isotonic lysis buffer containing 0.1% NP40. Co-immunoprecipitation was performed with Flag M2 affinity gel (Sigma Aldrich, A2220) and detected by Western Blot with Flag antibody (Sigma-Aldrich, F1804). On the same blot, the co-purified IVNS1ABP was detected by HA antibody (Biolegend, 901501).

### Induced pluripotent cells (iPSCs) and Neural progenitor cells (NPCs)

iPSCs were derived from primary cutaneous fibroblasts as previously described^87^. Fibroblasts of two patients (II.1 and II.4) and two healthy controls (I.1 and II.2) were reprogrammed using the CytoTune™-iPS 2.0 Sendai Reprogramming Kit (Thermo Fisher Scientific, A16517) in accordance with manufacturer’s instructions. Briefly, fibroblasts were transduced and after 7 days, were plated onto Matrigel Basement Membrane Matrix (Corning, 354234) in mTeSR1 medium (STEMCELL Technologies, 85850). iPSC colonies were picked between days 17-28 and maintained on Matrigel and mTeSR1 for expansion. NPCs were generated from iPSCs which were dissociated into single cells and seeded in low attachment U-bottom 96-well plate at a density of 10,000 cells/well in neural induction medium (DMEM/F12, Thermo Fisher Scientific, 10565-018) supplemented with B-27 (Thermo Fisher Scientific, 17504044), N-2 (Thermo Fisher Scientific, 17505048), 0.2 mM NEAA, 100 nM LDN 193189 (STEMCELL Technologies, 72148),

10 µM SB431542 (STEMCELL Technologies, 72234) and 10 µM ROCK Inhibitor Y-27632 (STEMCELL Technologies, 72304). After 6 days, cell aggregates were attached onto matrigel-coated plate and grown in neural expansion medium (DMEM/F12 (Thermo Fisher Scientific, 10565-018) supplemented with B-27, N-2, 0.2 mM NEAA and 20 ng/ml bFGF. After 3-6 days, rosette structures were manually cut and cultured in a neural expansion medium attached onto matrigel-coated plates^6^. Living cells were counted using Trypan Blue dye and glass hemocytometer.

### Fibroblast immortalization and CRISPR-Cas9-knockout

Primary fibroblasts were immortalized via pBABE NEO hTERT, human pCMV gag pol and human pCMV VSV-g retroviral infection using standard protocol (pBABE-neo largeTcDNA, Addgene plasmid 1780). CRISPR-Cas9 gene editing was carried out using a lentiviral infection, a co-editing and positive selection protocol published by the Doyon lab^88^. Cells were co-transfected with plasmids (Plasmid #52961) encoding guide RNAs against IVNS1ABP. Clonal selection was carried out by sequencing using a U6 sequencing primer. A diagnostic digestion using the *BamHI* and *EcoRV* enzymes was also performed to ensure that the selected colonies had the desired genomic alteration. LentiCRISPR-V2 and guide RNA were used to create stable deletions of *IVNS1ABP* and 2 µg/mL of puromycin was used for selection. The guide RNA target sequences for *IVNS1ABP* are Forward primer: 5’-CACCGACTGGGTGCAGCGTAGCATC-3’ and Reverse primer: 5’-AAACGATGCTACGCTGCACCCAGTC-3’.

### Neuron (NEUs) and motor neuron (MNs) differentiation from iPSCs

Neuron and motor neuron differentiation was carried out using CRISPR-Cas9 engineered isogenic iPSCs derived from patient II.1^54^. following previously established protocol^8^. In brief, human iPSCs were cultured in Neural Differentiation Medium (NDM), which is a mixture of DMEM/F12 and Neurobasal (1:1) and supplemented with 0.5x N-2, 0.5x B-27, and 1x Glutamax. To pattern the iPSCs to neural epithelial cells, SB431542 (2 μM), DMH-1 (2 μM), and CHIR99021 (3 μM) were added from day 1 to day 7. On day 7, the neural epithelial cells were split and continued to culture in NDM containing SB431542 (2 μM), DMH-1 (2 μM), CHIR99021 (3 μM), RA (0.1 μM), and SAG (1 μM) until day 14. At day 14, the motor neuron progenitors were lifted up for suspension culture in NDM containing RA (0.1 μM) and SAG (0.1 μM) until day 20. On day 20, the motor neuron progenitors were dissociated with TypLE and seeded onto Matrigel-coated plates. Cell markers, including HB9, Olig2, ChaT, and Tuj-1, were confirmed to characterize motor neuron identity. Lysotracker dye and live imaging were used to quantify lysosome motility, direction of movement, and velocity. Images were captured at 37°C with 5% CO2 (on-stage incubator and CO2 mixer (LCI)) using a Nikon Ti2 inverted microscope equipped with Yokogawa spinning disk confocal, GATACA super-resolution systems (SIM) and sCMOS camera (Prime 95B). Cells were imaged in multi channels by sequential laser excitations at 488 nm through a quad-bandpass dichroic mirror (Semrock) and single band emitters (Semrock). For lysotracker recording, movies were acquired on a single z plane with a speed of 2s/frame for 5 minutes, and exposure time was 100ms. Lysosome sizes were quantified with ImageJ macro ComDet.

### Aggresome staining

Fibroblasts were placed in 4-well chamber slides (Merck Millipore) with a seeding density between 9,000-12,000 cells per well. After 24 hours, cells were treated (12 hours) or not with 10 µM of MG132 (Merck/Calbiochem, 474790). Cells were fixed in 4% PFA for 30 minutes at room temperature, washed and permeabilized in 0.5% Triton X-100 for 15 minutes at room temperature. After three washes in PBS, cells were blocked in 1% BSA for 30 minutes at room temperature. Protein aggregates were stained using Proteostat® Aggresome Detection Reagent (Abcam, 139486, 1:1000 dilution) and nuclei were stained with Hoechst (Thermo Fisher Scientific, 62249). Slides were mounted in ProLong® Gold Antifade Reagent (Life Technologies) and images were taken with Brightfield and Widefield Zeiss AxioImager Z1 upright microscope.

### Ubiquitination assay and mass spectrometry (MS) analysis

Frozen patient and control NPCs were lysed in 8 M Urea in 50 mM Tris pH 8.0 lysis buffer. Cells were sonicated on ice. After centrifugation (40 min at 5000 rpm at 25°C). Protein concentration was measured using BCA kit (Thermo Fisher Scientific, 23225). Subsequently, samples were reduced with 20 mM with TCEP for 20 min at 25°C then alkylated with 55 mM CAA for 30 min at 25°C protected from direct light. The samples were then further diluted to Urea < 1M with 100 mM TEAB. Digestion performed with Lys-C 1:40 (enzyme:protein) overnight at 25°C followed by Trypsin 1:30 (enzyme:protein) overnight at 25°C Ubiquitin enrichment was performed using the PTMScan® Ubiquitin Remnant Motif (K-ε-GG) kit (Cell Signaling Technology, 5562) as per manufacturer’s instructions. The final desalting step of the protocol was performed on Oasis HLB 3cc 60 mg columns (Waters, WAT094226). Peptide quantification (Thermo 23290) was performed on the desalted samples to equalize sample amounts. The samples were dried and resuspended in 2% (v/v) Acetonitrile 0.5% (v/v) acetic acid , 0.06% (v/v) Trifluoroacetic acid (TFA) in water. The samples were separated by reverse phase liquid chromatography on the Easy nLC1000 coupled to Orbitrap Fusion™ Tribrid™ Mass Spectrometer with a 50 cm x 75 µm Easy Spray column (Thermo, ES803A). The samples were separated over a 120 min gradient with 0.1% formic acid in water (Merck, 1.59013.2500) and 99.9 % acetonitrile, 0.1% formic acid in water. The following acquisition parameters were applied: Data Dependent Acquisition/ positive mode with survey scan with 60000 Orbitap Analyzer (OT), scan range of 350-1250 m/z, and AGC target of 4e5; MS/MS collision induced dissociation in ion trap (IT) and AGC target of 1.5e4; Isolation window 1.2 m/z. Peak lists for subsequent searches were generated in Proteome Discoverer 2.2 (Thermo Fisher Scientific) using Mascot 2.6.1 (Matrix Science) and concatenated forward/decoy Human Uniprot database. The search parameters were as follow: MS precursor mass tolerance: 10 ppm; MS/MS fragment mass tolerance: 0.8 Da; 3 missed cleavages; static modifications: Carboamidomethyl (C); variable modifications: Oxidation (M), Deamidated (NQ), GG(K). False discovery rate estimation with 2 levels: Strict = FDR 1%, Medium = FDR 5%. Statistical and bioinformatics analysis: Precursor mass peak (MS1) intensities were quantified by label-free quantification (LFQ) and were normalized for total peptide abundance per condition. The enriched ubiquitinated peptide dataset was filtered for peptides identified with the GlyGlyK remnant motif. Proteins and peptides that were not identified in any of the 4 conditions were removed from further analyses. The abundances of ubiquitinated peptides were further normalized to the abundance of their respective unenriched protein. Differential analysis between the control and patient NPCs were performed on the normalized ubiquitinated peptides using two-sided unpaired Mann-Whitney test, generating the p-values and the average expression per condition.

### Amino acid starvation and drug treatment

NPCs were seeded in 6-well plates coated with matrigel and grown in a neural expansion medium. After 12 hours (70% confluent), medium was changed to 1) normal neural expansion medium, 2) amino acid-free neural expansion medium (US Biological #N1020, supplemented with B-27 and N-2) (starvation)^89^. To block the ubiquitin proteasome pathway and reduce proteolysis, half the wells cells were treated with 2.5 nM of Epoxomicin (Sigma Aldrich, E3652-50UG) for 4 and 8 hours. After 4 and 8 hours of incubation, harvested cells were kept frozen for protein extraction.

### Protein extraction and western blot

Cells were lysed and protein extracted using RIPA buffer (50 mM Tris HCl pH 7.5, 150 mM NaCl, 1% NP-40, 0.5% Na-Deoxycholate) supplemented with Complete Protease Inhibitor Cocktail tablets (EDTA free, Roche, 12245300). Protein extracts were quantified using Pierce BCA protein assay kit (Thermo Fisher Scientific, 23227) and 20 µg of protein lysate was loaded for western blotting. Protein extracts were then separated on 4–20% criterion TGX precast midi protein gel (Bio-Rad, 5678093) or 3-8% criterion XT Tris-Acetate Protein Gel (#3450129) with 1X XT tricine running buffer (#1610790), and transferred to PVDF membrane (Bio-rad, 170-4273) which was blocked in a 5% skim milk with TBST buffer. First antibodies were incubated overnight at 4°C and secondary antibodies at room temperature for 1 hour. All western blot results were confirmed in at least three fibroblast or neural progenitor cell subcultures derived from different controls and patients.

### Antibodies

For western blotting and cell immunostaining, all antibodies are commercially available and listed below:

| **Antibodies** | **Species** | **Company** | **Catalog #** | **Dilution** |
| --- | --- | --- | --- | --- |
| Ha | mouse | Biolegend | B901501 | 1:1000 |
| Flag | mouse | Sigma-Aldrich | F1804 | 1:1000 |
| Ubiquitin P4D1-A11 | mouse | Merck | 05-944 | 0.5 µg/ml |
| MAP1B | rabbit | Atlas Antibodies | HPA022275 | 1:1000 |
| MAP1B-LC (LC1) | mouse | Santa Cruz Biotechnology | SC-136472 | 1:200 |
| IVNS1ABP | rabbit | Atlas Antibodies | HPA003405 | 1:200 |
| Gigaxonin | rabbit | Atlas Antibodies | HPA049473 | 1:200 |
| P62 | mouse | BD Transduction | 610833 | 1:1000 |
| GAPDH | mouse | Santa Cruz Biotechnology | SC-32233 | 1:1000 |
| β-Catenin | rabbit | Sigma Aldrich | C2206 | 1:1000 |
| LAMP1 | mouse | Santa Cruz | sc-20011 | 1:500 |
| LAMP2 | rabbit | Abcam | ab18528 | 1:1000 |
| MNX1 | mouse | DSHB | 81.5C10-s | 1:50 |
| OLIG2 | rabbit | Merck | AB9610 | 1:500 |
| ChaT | goat | Merck | AB144P | 1:300 |
| MAP2 | rabbit | Abcam | 8707S | 1:1000 |
| p-NF | mouse | Sigma-Aldrich | MAB1592 | 1:1000 |
| Synaptotagmin 2 | mouse | DSHB | znp-1 | 1:1000 |

### Zebrafish CRISPR-Cas9 editing

Zebrafish were maintained and used according to the Singapore National Advisory Committee on Laboratory Animal Research Guidelines. Two guide RNAs against exon 4 of *Ivns1abpa* and *Ivns1abpb* were used with the following targeting sequences: 5′-CTGGAACCTCATGGTATCTC- 3′ and 5′-CAGGGTGGAGTTTCGCTGGT-3′ respectively. gRNAs were synthesized using the MEGAshortscript™ T7 Kit (Invitrogen, AM1354), treated with DNaseI and purified using RNeasy Mini Kit (Qiagen, 74104). 1 nl of a mixture containing 600-900pg of gRNA and 0.1 mg/ml of Cas9 protein was injected into the yolk of 1-cell AB zebrafish embryos. We selected, raised, and outbred two independent lines to eliminate potential off-target mutations. To verify mutations, we sanger sequenced injected embryos using PCR primers for *Ivns1abpa:* Forward: 5’- TTTTAATCCTGCAGCCAAGC-3’ and Reverse: 5’-AATTTGACCGCAACCAAGTC-3’, and for *Ivns1abpb*: Forward: 5’-TTTAATGAAACGAATTGTGCCTA-3’ and Reverse: 5’- GAATACACATCACTGACCAGACTTTT-3’.

### Maternal zygotic double knock-out fish (MZ dKO) generation

MZ dKO for i*vns1abpa* and *ivns1abpb* were generated by crossing homozygous zygotic single knock-out mutants for *ivns1abpa* and *ivns1abpb*, resulting in a heterozygous generation of mutants. Subsequently, the heterozygous mutants were incrossed and genotyped to identify the double knockout *ivns1abpa/b*.

Genotyping involved scaling zebrafish and overnight incubation at 65°C in a Proteinase K digestion solution, comprising Qiagen 10x PCR buffer, Qiagen Proteinase K (20 mg/ml), and Molecular Grade Water (SH30538.03). After centrifugation, the samples were used for Polymerase Chain Reaction with Hot Staq PCR Master mix (#203203), using 18 µl of PCR mix and 2 µl of DNA from digested zebrafish scales. Sanger sequencing was then carried out using a Big Dye mastermix with forward and reverse primers. Once the genotype was confirmed, the homozygous double knockout embryos were incrossed to obtain maternal-zygotic ivns1abpa and ivns1abpb double knock-out fish, and checked by Sanger sequencing.

### Zebrafish reverse transcription quantitative PCR (RT-qPCR)

Embryos were harvested and de-chrorionated at different stages of development. About 20-30 embryos were pooled and lysed using 600 µl of BME-RLT buffer together with a 3 ml syringe and BD precision GlideTM 23G 114 TW to homogenize it. Lysates were loaded into a QIA Shredder column (Qiagen, 79656) to reduce viscosity. Columns were then centrifuged at 10,000 rpm for 2 mins. Supernatants were collected for RNA extraction using RNeasy Mini Kit (Qiagen, 74104). 1 µg of the extracted RNA were converted to cDNA using the Iscript Reverse Transcription Supermix (Bio-Rad, 1708840). RT-qPCR reactions were carried out using the Power SYBR Green PCR Master Mix (Applied Biosystems, 4309155). Three technical replicates per sample were done in three biological replicates. *Actin* was used as the housekeeping gene. Data were analyzed and compared using Student’s independent t-test using GraphPad Prism (GraphPad Software, La Jolla, California, USA). Data are plotted in column graphs with standard error of the mean (SEM). Statistical significance was assumed with a p-value <0.001 (indicated by ***).

### Zebrafish whole mount in situ hybridization (WISH)

WISH was carried out using DIG-labeled and fluorescein-labeled antisense probes as previously described^90^. Sense and antisense probes for *ivns1abpa* and *ivns1abpb* were synthesized with Sp6 and T7 RNA polymerases from a linearized pGEMT-easy vector containing *ivns1abpa* and *ivns1abpb* PCR inserts. Fluorescein-labeled probes were detected using P-iodonitrotetrazolium violet plus 5-bromo-4-chloro-3-indolyl phosphate (INT/BCIP) and DIG-labeled probes with nitro blue tetrazolium (NBT). After staining, embryos were fixed in 4% paraformaldehyde and submerged in 70% glycerol for imaging using Leica Application Suite (Leica Microsystems Switzerland Limited).

### Zebrafish western blot

40 embryos were dechorionated 1 day post fertilization (dpf) and deyolked using Ginzburg Fish Ringer (6.5 g NaCl; 0.25 g KCl; 0.3 g CaCl2; 0.2 g NaH CO3 in 1 liter ddH2O). Embryos were then disrupted using a syringe and left on a thermal shaker for 5 minutes at 1,100 rpm. Lysed embryos were centrifuged at 300 g for 30 seconds. The pellet was washed 2 times in 1 ml of wash buffer (110 mM NaCl; 3.5 mM KCl; 2.7 mM CaCl2; 10 mM Tris/Cl (pH8.5)), then left on shaker for 2 minutes at 1,100 rpm. Pellets were then centrifuged and lysed in RIPA buffer (250 mM Tris, pH: 7.5; 150 mM NaCl; 1% NP-40; 0.5% Na deoxycholate) together with Protease Inhibitor (Sigma Aldrich, S8820). Pellets were kept on ice for 20 minutes and centrifuged at 13,000 rpm for 20 minutes. Supernatant was loaded and immunoblotting was performed using anti-IVNS1ABP antibodies.

### Zebrafish primary motor neuron axon staining and quantification

Embryos were collected 1 dpf, dechorionated, and fixed 31 hours post fertilization (hpf) in 4% paraformaldehyde at 4°C overnight on a rocker. Fixed embryos were then washed 3 times 5 minutes in PBST, followed by 5 minutes wash in water, 4 hours incubation in water on a rocker, 1 hour incubation in 500 µl PBDT. Embryos were then incubated with primary antibody α- synaptotagmin 2 (DSHB #ZNP-1) diluted in PBDT overnight at 4°C. The day after, embryos were washed 4 times, with 1 hour interval, in PBST with 0.1% TritonX-100 on rocker at room temperature. Embryos were then incubated with a secondary antibody biotin-coupled anti mouse for 1:1000 (VECTASTAIN® Elite ABC-HRP Kit, Peroxidase Mouse IgG, PK-6102) in PBST at 4°C overnight. Secondary antibody was then discarded and embryos were washed 4 times, with 1 hour interval. Embryos were then incubated in Vectastin ABC solution for 1 hour at room temperature on a rocker, then washed 4 times, with 30 minutes interval, in PBST with 0.1% TritonX-100, at room temperature on rocker. Embryos were then incubated for 30 minutes at room temperature in DAB solution (SIGMAFAST™ 3,3′-Diaminobenzidine tablets, Sigma Aldrich, D4418), then transferred in a mix of DAB and urea for 2 minutes, then washed in PBST 3 times, with 5 minutes interval. Stained embryos were stored at 4°C in PBST and visualized using Nikon Ni-E upright brightfield & widefield microscope. Analysis was done using ImageJ and NIS-Elements Viewer 5.21. For quantification, the phenotype of the axonal projections of primary motor neurons were analyzed specifically in hemisegments 2-6 (from anterior to posterior). To avoid experimenter bias, a blind analysis was performed. Embryos were individually assessed on their axonal projections without knowing embryos’ genotype. The folder names for each genotype were relabeled using an R program. Consequently, all folders were rendered anonymous, preventing the identification of their respective genotypes. Three types of axons were then categorized: normal axons, where all axons projected downwards in an orderly manner; poorly fasciculated axons, displaying less organized downward projection; and aberrant axon pathfinding, characterized by projections into neighboring somites. Subsequently, the scores were averaged and quantified using Estimation Stats (https://www.estimationstats.com/#/analyze/two-independent-groups).

### Zebrafish locomotion swim assay

Zebrafish behavioral experiments were conducted according to approved protocols (A*STAR IACUC #191501 and #231797). The swimming behavior of a total of 139 zebrafish larvae (72 control and 69 MZ dKO ) at 7 dpf was recorded on a custom-designed hardware via a Basler Ace (acA1300-200um; 1,280 × 1,024) camera with a 25 mm lens attachment placed 65 cm above a 24-well plate. Each well was coated with 1 ml of 2% agarose and backlit by uniform white LED lightbox. Using a dropper, larvae were placed singly into each well and acclimated for 5 minutes, followed by 2-minute video recording larval movement. Such videos of 2 minutes were acquired with 3 minutes rest between each video. 5 mM of pentylenetetrazole (PTZ, Sigma, 54-95-5) was added to each well after the first video. Medium- and high-speed swim bouts were defined as 8 to 16 mm/sec and >16 mm/sec respectively. Tracks, distance traveled, velocity, and immobility frequency were quantified automatically using custom-written scripts in Python. Repository for pull requests: https://github.com/mechunderlyingbehavior/24-Well-Larval-Locomotion.git. Statistical analysis was performed using Estimation Statistics. Two-sided permutation t-test was used for p-values presented^91^.

---

Supplemental Figure 1. Mutations in *IVNS1ABP* and *GAN* cause syndromes with similar clinical manifestations

**(A)** Screenshots demonstrating steppage gait and difficulty to walk of the three patients present with progressive and severe neuropathy.

**(B)** Body photograph of patients II.4 and II.5 displaying hyperpigmented macules.

**(C)** Variant filtering identified a single germline biallelic deleterious variant in the *IVNS1ABP* gene.

**(D)** AlphaFold3 prediction of the binding of DAPK1 peptide, containing a hydrophobic LPDLV motif, with Leu1339 interacting with KLHL20 Klech domain. IVNS1ABP is also predicted to bind to KLHL20 in a similar manner as DAPK1. See also Figure 1H.

**(E)** Photographs of a *GAN*-null patient displaying spastic paraplegia with severe muscle atrophy, scoliosis, club foot, and kinky hair and eyebrows. He has reticular skin hyperpigmentation on the axillary area, but the hyperpigmented skin macules on the torso are mainly due to healing scabies wounds.

Supplemental Figure 2. Altered IVNS1ABP induces protein aggregation in patients’ cells

**(A)** Proteostat® staining shows significant increase of protein aggregates in two patients’ iPSCs (II.1 and II-4) compared to unaffected control (I.1) and non-carrier control’s iPSCs (II.2). Scale bar = 50 µm.

**(B)** Immunostaining of lysosome-associated membrane proteins 1 and 2 (LAMP1, LAMP2) shows significant increase of the lysosomal network in two patients’ iPSCs (II.1 and II.4) compared to unaffected control (I.1) and non-carrier control’s iPSCs (II.2). Scale bar = 50 µm.

Supplemental Figure 3. Protein ubiquitination is altered in IVNS1ABP mutant cells Immunostaining and quantification of Vimentin in patient cells (II.1) (n=297) show increased signal compared to healthy control (II.2) (n=240), validating our approach. Scale bar = 10 µm. Error bars indicate mean ± SEM. Unpaired t test, **** p < 0.0001.

Supplemental Figure 4. Altered IVNS1ABP induces enlarged lysosomal network in patients’ cells

Immunostaining and quantification of OLIG2 and MNX1 show that isogenic iPSCs differentiated into motor neuron progenitors with no significant difference (ns, unpaired t test).

Supplemental Figure 5. Ivns1abpa and *ivns1abpb* paralogs in zebrafish

**(A)** qRT-PCR shows the expression level of *ivns1abpa* and *ivns1abpb* transcripts at different stages of zebrafish embryo development, from zygotic to 5 dpf.

**(B)** WISH shows the spatio-temporal expression of *ivns1abpa* and *ivns1abpb* at different stages of zebrafish embryo development, from zygotic to 5 dpf.

**(C)** Phylogeny and synteny analysis show that *ivns1abpa* is closer to human *IVNS1ABP* than *ivns1abpb*.

Supplemental Figure 6. Generation and characterization of *Ivns1abpa* and *Ivns1abpb* maternal zygotic double knockout zebrafish

**(A)** Schematic representation of CRISPR-Cas9 engineered mutations generated in zebrafish

*ivns1abpa* and *ivns1abpb*.

**(B)** CRISPR-Cas9 engineered mutations were confirmed by Sanger sequencing in *ivns1abpa* and

*ivns1abpb* maternal zygotic double knockout zebrafish (MZ dKO).

**(C)** Western blot of overexpressed zebrafish *ivns1abpa* and *ivns1abpb* shows that IVNS1ABP antibodies recognized zebrafish Ivns1abp proteins. Two independent frameshift mutations in *ivns1abpa* induced almost complete loss of ivns1abp proteins in 1 dpf zebrafish embryos.

**(D)** Compared to WT (n=72), 7 dpf MZ dKO (n=67) larvae show significant differences in locomotor behaviors, including a lower number of inter-swimming bout pauses and intermediate- speed bouts where larvae swam 8-16 mm/s. Larvae were challenged with 5 mM of Pentylenetetrazol (PTZ) for 5, 10 and 15 minutes (mins). In Gardener-Altman and Cummings estimation plots, the mean difference between the two groups (black dot) and 95% confidence intervals (black bar) are plotted on the secondary axis. The orange-shaded area in orange shows the 5000 bootstrapped confidence intervals. Two-sided permutation t-test for p-values are represented as * p < 0.05, *** p < 0.001, **** p < 0.0001.

Supplemental Table 1. Protein ubiquitination is altered in IVNS1ABP mutant cells

Lists of top ubiquitinated proteins, peptides and targets.

## Notes

### Competing Interest Statement

The authors have declared no competing interest.

### Funding Statement

This study was funded by multiple sources: NMRC Open Fund - Young Individual Research Grant (OF-YIRG/0048/2017); NMRC Open Fund - Young Individual Research Grant (OF-YIRG22jul-0021); Duke-NUS-KPFA/2020/0038; Singapore Ministry of Education, MOE2018-T2-2-103; Singapore Ministry of Health, MOH-000207 and 000212; Singapore Ministry of Education MOE-T2EP30220-0020; China Scholarship Council - Nuffield Department of Medicine Scholarship; Wellcome (106169/ZZ14/Z); Strategic Positioning Fund for Genetic Orphan Diseases (SPF2012/005)

### Author Declarations

The institutional review board of the Agency for Science, Technology and Research (A*STAR) gave ethical approval for this work, under IRB number 2019-087.

