## Supplemental Data for "Loss of IVNS1ABP, a gigaxonin paralogue, leads to a progeroid neuropathy due to impaired proteostasis"

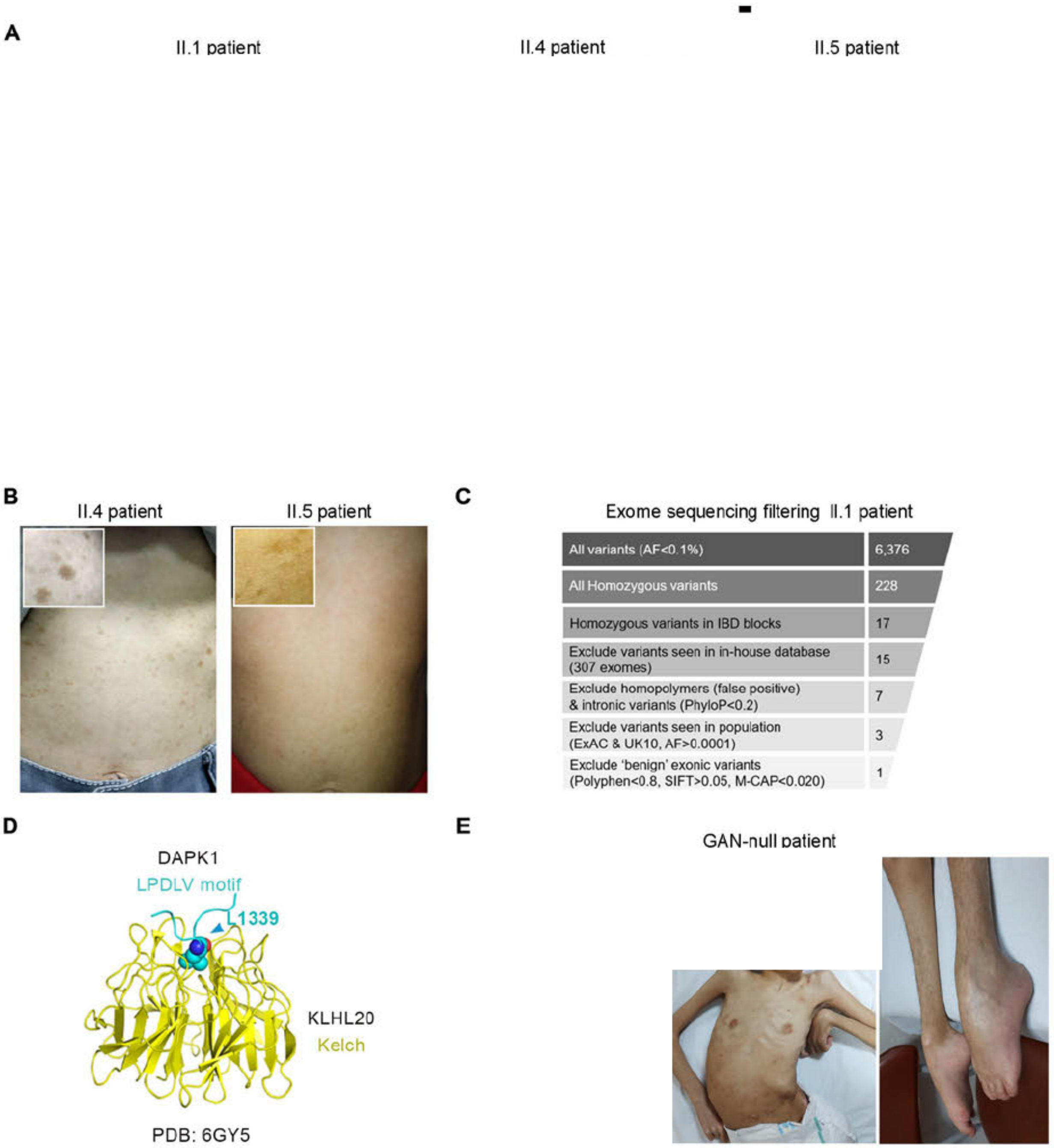

Figure S1
C. Bonnard *et al.*, (2024)

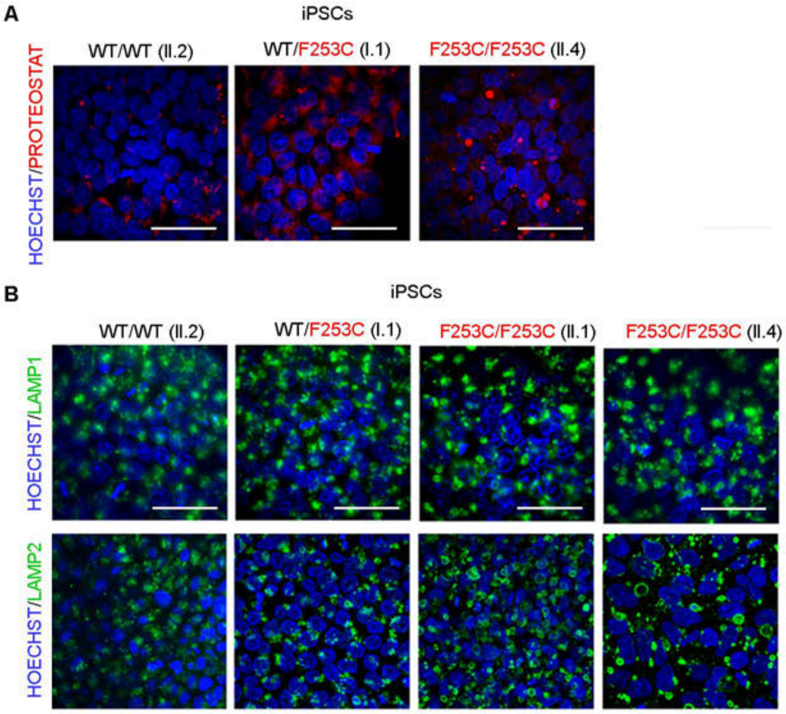

Figure S2
C. Bonnard *et al.*, (2024)

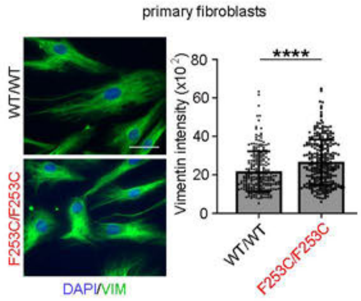

Figure S3
C. Bonnard *et al.*, (2024)

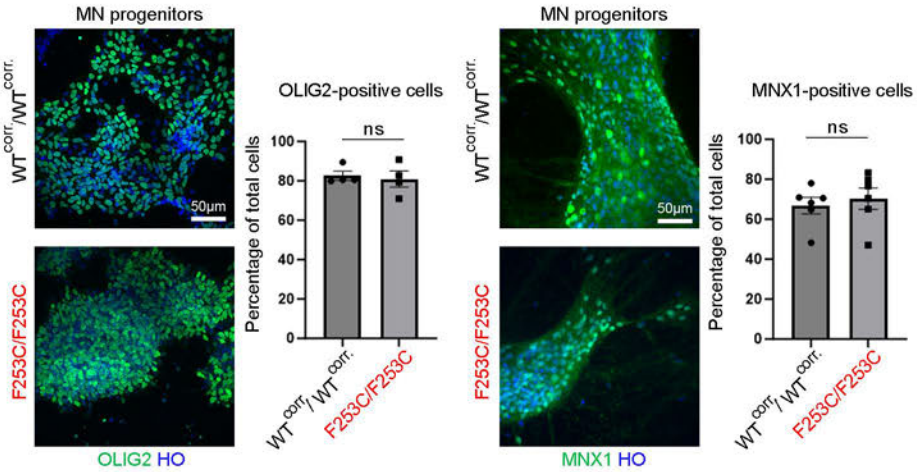

Figure S4
C. Bonnard *et al.*, (2024)

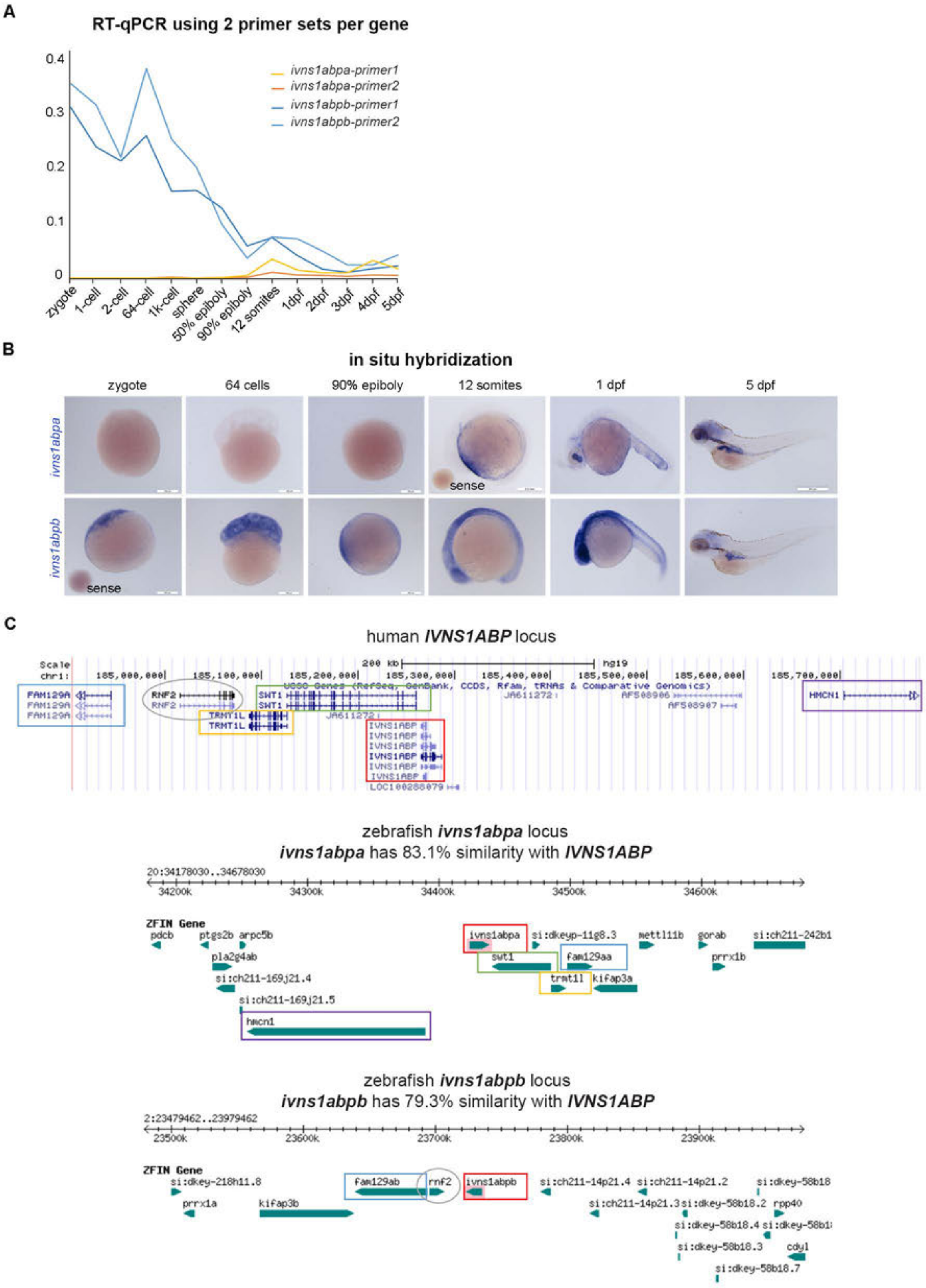

Figure S5
C.Bonnard *et al.*, (2024)

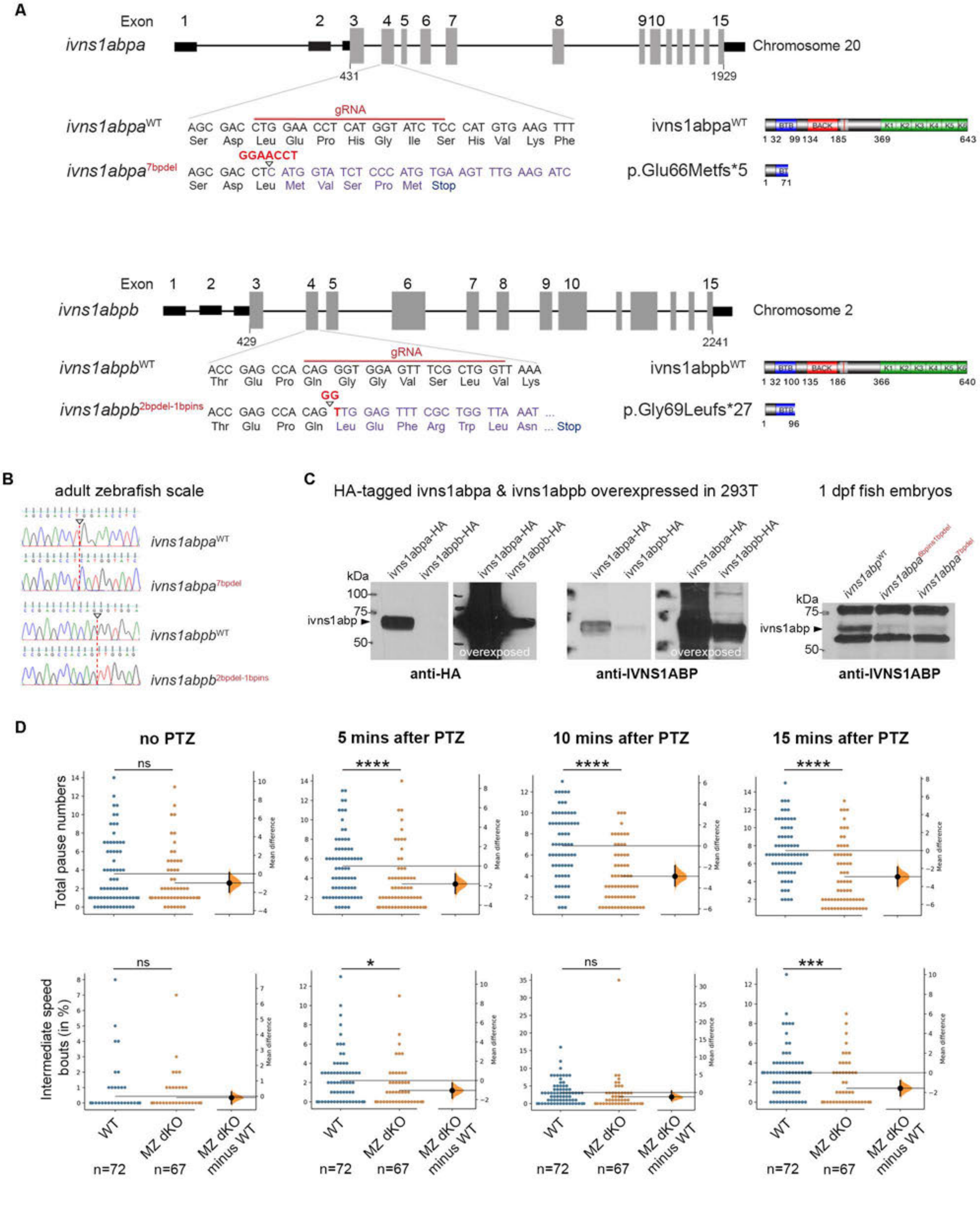

Figure S6
C.Bonnard *et al.*, (2024)

| | Top list of ubiquitinated proteins with significant fold change (logFC) in two homozygous (HMZ) patients (II.1 & II.4) versus two controls wildtype (WT II.2) and heterozygous (HTZ I.1) | | | | | | | | |
|---|---|---|---|---|---|---|---|---|---|
| | Protein name | *Gene* | II.2 (WT) | I.1 (HTZ) | II.1 (HMZ) | II.4 (HMZ) | p value | WTav | MTav |
| 1 | Alpha-enolase | *ENO1* | 253,738,039 | 233,746,418 | 167,852,203 | 189,183,389 | 0.0471 | 243,742,229 | 178,517,796 |
| 2 | Glucosidase 2 subunit beta | *PRKCSH* | 36,156,970 | 37,294,114 | 44,963,055 | 44,837,468 | 0.0419 | 36,725,542 | 44,900,261 |
| 3 | 60S ribosomal protein L5 | *RPL5* | 22,565,294 | 22,109,299 | 27,170,165 | 26,581,446 | 0.0083 | 22,337,297 | 26,875,806 |
| 4 | Heterogeneous nuclear ribonucleoprotein H2 | *HNRNPH2* | 3,177,915 | 3,474,061 | 2,172,992 | 1,779,883 | 0.0371 | 3,325,988 | 1,976,437 |
| 5 | Serine/arginine-rich splicing factor 6 | *SRSF6* | 4,675,644 | 4,815,498 | 5,136,883 | 5,295,910 | 0.0485 | 4,745,571 | 5,216,396 |
| 6 | Isoform 8 of Protein transport protein Sec31A | *SEC31A* | 5,005,881 | 5,101,888 | 6,811,001 | 6,819,738 | 0.0165 | 5,053,884 | 6,815,369 |
| 7 | 40S ribosomal protein S23 | *RPS23* | 7,030,710 | 6,489,940 | 9,079,727 | 8,411,920 | 0.0474 | 6,760,325 | 8,745,824 |
| 8 | UMP-CMP kinase | *CMPK1* | 5,188,959 | 5,498,722 | 3,706,928 | 3,489,043 | 0.0161 | 5,343,840 | 3,597,986 |
| 9 | Methylmalonate-semialdehyde dehydrogenase [acylating], mitochondrial | *ALDH6A1* | 1,009,509 | 1,225,089 | 5,224,426 | 4,687,541 | 0.0232 | 1,117,299 | 4,955,983 |
| 10 | Regulation of nuclear pre-mRNA domain-containing protein 1B | *RPRD1B* | 5,486,370 | 4,848,454 | 2,924,887 | 2,454,401 | 0.0301 | 5,167,412 | 2,689,644 |
| 11 | Proteasome activator complex subunit 2 | *PSME2* | 6,574,865 | 6,759,166 | 7,628,436 | 7,827,150 | 0.0162 | 6,667,016 | 7,727,793 |
| 12 | Exportin-7 | *XPO7* | 1,885,534 | 1,571,306 | 3,003,602 | 2,657,977 | 0.0428 | 1,728,420 | 2,830,789 |
| 13 | Double-stranded RNA-binding protein Staufen homolog 2 | *STAU2* | 826,243 | 807,304 | 920,381 | 953,286 | 0.0406 | 816,773 | 936,834 |
| 14 | Isoform 2 of AP-1 complex subunit gamma-1 | *AP1G1* | 2,177,996 | 2,713,351 | 3,953,431 | 4,384,684 | 0.0411 | 2,445,673 | 4,169,057 |
| 15 | Thioredoxin reductase 1, cytoplasmic | *TXNRD1* | 6,360,014 | 6,949,799 | 10,597,122 | 10,462,265 | 0.0389 | 6,654,906 | 10,529,693 |
| 16 | Nuclear pore glycoprotein p62 | *NUP62* | 194,035 | 312,774 | 985,746 | 897,002 | 0.0145 | 253,405 | 941,374 |
| 17 | Uridine 5'-monophosphate synthase | *UMPS* | 2,643,323 | 3,254,054 | 5,794,254 | 4,984,072 | 0.0468 | 2,948,688 | 5,389,163 |
| 18 | Prolyl endopeptidase | *PREP* | 1,424,477 | 1,582,483 | 655,172 | 874,623 | 0.0392 | 1,503,480 | 764,898 |
| 19 | Isoform 2 of Cystathionine beta-synthase | *CBS* | 1,239,382 | 762,118 | 2,664,919 | 3,176,507 | 0.0320 | 1,000,750 | 2,920,713 |
| 20 | Calcium-binding protein 39 | *CAB39* | 1,181,102 | 1,273,196 | 599,742 | 464,311 | 0.0196 | 1,227,149 | 532,026 |
| 21 | Mini-chromosome maintenance complex-binding protein | *MCMBP* | 173,211 | 291,392 | 751,242 | 860,679 | 0.0194 | 232,302 | 805,961 |
| 22 | Costars family protein ABRACL | *ABRACL* | 411,472 | 439,530 | 264,889 | 248,893 | 0.0188 | 425,501 | 256,891 |
| 23 | Nuclear receptor-binding protein | *NRBP1* | 244,697 | 209,813 | 64,231 | 101,227 | 0.0298 | 227,255 | 82,729 |
| 24 | Mitochondrial carrier homolog 2 | *MTCH2* | 503,657 | 361,894 | 1,356,402 | 1,621,239 | 0.0381 | 432,776 | 1,488,821 |
| 25 | Ras-related GTP-binding protein D | *RRAGD* | 395,518 | 362,456 | 257,101 | 282,532 | 0.0395 | 378,987 | 269,816 |
| 26 | Isoform 2 of Peptidyl-prolyl cis-trans isomerase FKBP8 | *FKBP8* | 222,266 | 207,138 | 265,756 | 284,656 | 0.0415 | 214,702 | 275,206 |
| 27 | RNA polymerase II elongation factor ELL3 | *ELL3* | 738,435 | 750,141 | 475,555 | 441,068 | 0.0230 | 744,288 | 458,312 |

**Top list of ubiquitinated peptides with significant fold change (logFC) in two homozygous (HMZ) patients (II.1 & II.4) versus two controls wildtype (WT II.2) and heterozygous (HTZ I.1)**

| | Uniprot id | protein name | *Gene* | II.2 (WT) | I.1 (HTZ) | II.1 (HMZ) | II.4 (HMZ) | p value | WTav | MTav |
|---|---|---|---|---|---|---|---|---|---|---|
| 1 | Q15004 [24-41] 1D[N11]1GG[K1] | PCNA-associated factor | *PCLAF* | 92,345 | 81,620 | 350,598 | 361,567 | 0.0008 | 86,982 | 356,082 |
| 2 | O43175 [381-394] 1D[N12]1GG[K4] | D-3-phosphoglycerate dehydrogenase | *PHGDH* | 66,489 | 57,045 | 178,801 | 167,317 | 0.0051 | 61,767 | 173,059 |
| 3 | Q9ULK5 [372-382] 1GG[K8] | Vang-like protein 2 | *VANGL2* | 13,944 | 41,303 | 838,698 | 777,508 | 0.0092 | 27,624 | 808,103 |
| 4 | P13667 [235-246] 1GG[K11] | Protein disulfide-isomerase A4 | *PDIA4* | 14,480 | 19,205 | 46,496 | 42,471 | 0.0134 | 16,843 | 44,484 |
| 5 | P20700 [529-542] 1GG[K4] | Lamin-B1 | *LMNB1* | 20,378 | 26,238 | 54,291 | 49,773 | 0.0193 | 23,308 | 52,032 |
| 6 | P68104 [393-408] 1GG[K3] | Elongation factor 1-alpha 1 | *EEF1A1* | 39,961 | 31,584 | 69,954 | 76,259 | 0.0232 | 35,772 | 73,107 |
| 7 | Q8N0X7 [460-475] 1GG[K6] | Spartin | *SPART* | 48,172 | 66,184 | 135,506 | 121,283 | 0.0283 | 57,178 | 128,394 |
| 8 | P68036-3 [181-191] 1GG[K9] | Isoform 3 of Ubiquitin-conjugating enzyme E2 L3 | ***UBE2L3*** | 145,488 | 119,269 | 262,646 | 299,609 | 0.0289 | 132,379 | 281,128 |
| 9 | P62913 [170-178] 1GG[K9] | 60S ribosomal protein L11 | *RPL11* | 63,366 | 57,250 | 102,097 | 103,962 | 0.0309 | 60,308 | 103,030 |
| 10 | Q13286 [256-272] 1GG[K7] | Battenin | *CLN3* | 110,190 | 119,520 | 428,711 | 390,306 | 0.0319 | 114,855 | 409,508 |
| 11 | P61088 [71-82] 1GG[K4] | Ubiquitin-conjugating enzyme E2 N | *UBE2N* | 310,585 | 292,777 | 381,802 | 372,069 | 0.0341 | 301,681 | 376,935 |
| 12 | P68036-3 [130-140] 1GG[K2] | Isoform 3 of Ubiquitin-conjugating enzyme E2 L3 | ***UBE2L3*** | 367,511 | 511,829 | 899,490 | 1,001,458 | 0.0363 | 439,670 | 950,474 |
| 13 | P45973 [144-159] 1GG[K] | Chromobox protein homolog 5 | *CBX5* | 181,323 | 194,661 | 133,568 | 127,051 | 0.0371 | 187,992 | 130,310 |
| 14 | P11142 [518-526]; P0DMV8 [518-526] 1O[M1]1GG[K7] | Heat shock cognate 71 kDa protein | *HSPA8* | 77,995 | 83,296 | 244,315 | 219,601 | 0.0431 | 80,646 | 231,958 |
| 15 | Q7Z569 [377-400] 2C[C15C20]2GG[K5K9] | BRCA1-associated protein | *BRAP* | 61,446 | 45,855 | 120,330 | 145,057 | 0.0465 | 53,650 | 132,694 |
| 16 | P62714 [37-49]; P67775 [37-49] 1GG[K5] | Serine/threonine-protein phosphatase 2A catalytic subunit beta isoform | *PPP2CB* | 99,913 | 97,869 | 198,207 | 183,564 | 0.0466 | 98,891 | 190,885 |
| 17 | P11142 [518-526]; P0DMV8 [518-526] 1GG[K7] | Heat shock cognate 71 kDa protein | *HSPA8* | 430,899 | 407,282 | 780,296 | 862,300 | 0.0483 | 419,090 | 821,298 |

**Top list of ubiquitinated peptide targets matched against fold change (protein_fc) and p-value (protein.Pvalue) in corresponding protein.**

| | Uniprot id | protein name | *Gene* | II.2 (WT) | I.1 (HTZ) | II.1 (HMZ) | II.4 (HMZ) | pvalue | WTav | MTav |
|---|---|---|---|---|---|---|---|---|---|---|
| 1 | Q99426 [197-211] 1D[N13]1GG[K7] | Tubulin-folding cofactor B | *TBCB* | 0.0014 | 0.0013 | 0.0020 | 0.0020 | 0.0064 | 0.0014 | 0.0020 |
| 2 | P63104 [139-157] 1GG[K1] | 14-3-3 protein zeta/delta | *YWHAZ* | 0.0002 | 0.0002 | 0.0004 | 0.0004 | 0.0143 | 0.0002 | 0.0004 |
| 3 | P15121 [257-269] 1GG[K7] | Aldose reductase | *AKR1B1* | 0.0021 | 0.0023 | 0.0031 | 0.0032 | 0.0239 | 0.0022 | 0.0032 |
| 4 | P62258 [62-73] 1GG[K8] | 14-3-3 protein epsilon | *YWHAE* | 0.0004 | 0.0003 | 0.0006 | 0.0007 | 0.0275 | 0.0003 | 0.0006 |
| 5 | Q15233 [97-107] 1GG[K3] | Non-POU domain-containing octamer-binding protein | *NONO* | 0.0004 | 0.0003 | 0.0007 | 0.0006 | 0.0281 | 0.0004 | 0.0007 |
| 6 | Q96FW1 [84-94] 1C[C8]1GG[K1] | Ubiquitin thioesterase OTUB1 | *OTUB1* | 0.0018 | 0.0026 | 0.0053 | 0.0059 | 0.0321 | 0.0022 | 0.0056 |
| 7 | P84103 [78-86] 1D[N5]1GG[K8] | Serine/arginine-rich splicing factor 3 | *SRSF3* | 0.0047 | 0.0036 | 0.0086 | 0.0080 | 0.0389 | 0.0041 | 0.0083 |
| 8 | P68036-3 [130-140] 1GG[K2] | Isoform 3 of Ubiquitin-conjugating enzyme E2 L3 | *UBE2L3* | 0.0255 | 0.0387 | 0.0798 | 0.0967 | 0.0390 | 0.0321 | 0.0882 |
| 9 | Q06830 [169-190] 1C[C5]1GG[K10] | Peroxiredoxin-1 | *PRDX1* | 0.0003 | 0.0004 | 0.0006 | 0.0007 | 0.0394 | 0.0003 | 0.0007 |
| 10 | P08670 [441-450] 1GG[K5] | Vimentin | *VIM* | 0.0000 | 0.0001 | 0.0001 | 0.0001 | 0.0481 | 0.0001 | 9.5469 |

**Top 20 pathways enrichment analysis with the ubipeptide results**

| ID | Description | setSize | enrichmentScore | NES | p value | p.adjust | q values | rank | leading_edge |
|---|---|---|---|---|---|---|---|---|---|
| R-HSA-168256 | Immune System | 81 | 0.394677963 | 1.9579749 | 0.0010152 | 0.0044235 | 0.0024932 | 298 | tags=77%, list=49%, signal=45% |
| R-HSA-1266738 | Developmental Biology | 52 | 0.456079873 | 2.0903467 | 0.0010331 | 0.0044235 | 0.0024932 | 149 | tags=52%, list=25%, signal=43% |
| R-HSA-8953854 | Metabolism of RNA | 55 | 0.43914376 | 2.0330473 | 0.0010341 | 0.0044235 | 0.0024932 | 106 | tags=42%, list=17%, signal=38% |
| R-HSA-168249 | Innate Immune System | 53 | 0.422965382 | 1.9371094 | 0.0010363 | 0.0044235 | 0.0024932 | 298 | tags=81%, list=49%, signal=45% |
| R-HSA-422475 | Axon guidance | 45 | 0.526296791 | 2.3517864 | 0.0010428 | 0.0044235 | 0.0024932 | 149 | tags=60%, list=25%, signal=49% |
| R-HSA-5663205 | Infectious disease | 40 | 0.470011733 | 2.0467143 | 0.0010571 | 0.0044235 | 0.0024932 | 83 | tags=40%, list=14%, signal=37% |
| R-HSA-5653656 | Vesicle-mediated transport | 33 | 0.426037513 | 1.7922912 | 0.001065 | 0.0044235 | 0.0024932 | 253 | tags=73%, list=42%, signal=45% |
| R-HSA-199991 | Membrane Trafficking | 30 | 0.458408656 | 1.8765067 | 0.0010834 | 0.0044235 | 0.0024932 | 253 | tags=77%, list=42%, signal=47% |
| R-HSA-194315 | Signaling by Rho GTPases | 30 | 0.48177795 | 1.9721694 | 0.0010834 | 0.0044235 | 0.0024932 | 145 | tags=57%, list=24%, signal=45% |
| R-HSA-195258 | RHO GTPase Effectors | 29 | 0.494899788 | 2.0140785 | 0.0010846 | 0.0044235 | 0.0024932 | 145 | tags=59%, list=24%, signal=47% |
| R-HSA-71291 | Metabolism of amino acids and derivatives | 28 | 0.515457693 | 2.0790955 | 0.0010893 | 0.0044235 | 0.0024932 | 99 | tags=50%, list=16%, signal=44% |
| R-HSA-376176 | Signaling by ROBO receptors | 28 | 0.538835138 | 2.1733883 | 0.0010893 | 0.0044235 | 0.0024932 | 142 | tags=61%, list=23%, signal=49% |
| R-HSA-72766 | Translation | 26 | 0.504990508 | 2.0053614 | 0.0010965 | 0.0044235 | 0.0024932 | 103 | tags=50%, list=17%, signal=43% |
| R-HSA-168254 | Influenza Infection | 24 | 0.581865839 | 2.2590866 | 0.0011123 | 0.0044235 | 0.0024932 | 99 | tags=54%, list=16%, signal=47% |
| R-HSA-168255 | Influenza Life Cycle | 24 | 0.581865839 | 2.2590866 | 0.0011123 | 0.0044235 | 0.0024932 | 99 | tags=54%, list=16%, signal=47% |
| R-HSA-9010553 | Regulation of expression of SLITs and ROBOs | 24 | 0.56164222 | 2.1805686 | 0.0011123 | 0.0044235 | 0.0024932 | 99 | tags=54%, list=16%, signal=47% |
| R-HSA-1799339 | SRP-dependent cotranslational protein targeting to membrane | 22 | 0.593006086 | 2.246981 | 0.0011236 | 0.0044235 | 0.0024932 | 103 | tags=59%, list=17%, signal=51% |
| R-HSA-927802 | Nonsense-Mediated Decay (NMD) | 22 | 0.606081531 | 2.2965257 | 0.0011236 | 0.0044235 | 0.0024932 | 99 | tags=59%, list=16%, signal=51% |
| R-HSA-975957 | Nonsense Mediated Decay (NMD) enhanced by the Exon Junct | 22 | 0.606081531 | 2.2965257 | 0.0011236 | 0.0044235 | 0.0024932 | 99 | tags=59%, list=16%, signal=51% |
| R-HSA-6791226 | Major pathway of rRNA processing in the nucleolus and cytoso | 21 | 0.583865908 | 2.1791561 | 0.0011338 | 0.0044235 | 0.0024932 | 99 | tags=57%, list=16%, signal=50% |

### Table S1

#### C. Bonnard *et al.*, (2024)
